# Benchmarking ten frontier large language models on 1,477 board-style multiple choice questions in hematology

**DOI:** 10.64898/2026.09.01.26361881

**Authors:** Martina Radoynova, Mohamed Benouis, Freya Schulze, Martin MK Schneider, Susann Winter, Martin Bornhäuser, Jan Moritz Middeke, Jan-Niklas Eckardt

**Affiliations:** Department of Internal Medicine I, University Hospital Carl Gustav Carus, TUD Dresden University of Technology, Dresden, Germany; Else Kröner Fresenius Center for Digital Health, TUD Dresden University of Technology, Dresden, Germany; German Cancer Consortium (DKTK), Partner Site Dresden, and German Cancer Research Center (DKFZ), Heidelberg, Germany; National Center for Tumor Diseases (NCT), NCT/UCC Dresden, a partnership between DKFZ, Faculty of Medicine and University Hospital Carl Gustav Carus, TUD Dresden University of Technology, and Helmholtz-Zentrum Dresden-Rossendorf (HZDR), Germany; Mildred Scheel Early Career Center, National Center for Tumor Diseases (NCT/UCC) Dresden, Faculty of Medicine and University Hospital Carl Gustav Carus, TUD Dresden University of Technology, Dresden, Germany

**Author notes:** **Correspondence:** Jan-Niklas Eckardt, MD, MSc, MHBA; Department of Internal Medicine I, University Hospital Carl Gustav Carus, Technical University Dresden and Else-Kröner-Fresenius Center for Digital Health, Technical University Dresden, Fetscherstraße 74, 01307 Dresden Germany.

**Keywords:** artificial intelligence, large language models, benchmark

## Abstract

Large Language Models (LLMs) are increasingly used by clinicians and patients for medical queries, yet their accuracy and safety at the specialist level in hematology remain insufficiently characterised. We benchmarked ten frontier proprietary and open-weight LLMs across two generations on 1,477 board-style hematology multiple-choice questions (MCQs) derived from five educational datasets spanning nine disease areas and six clinical skill domains, including text-only and multimodal case vignettes. Claude Opus 5 had the highest mean accuracy (92.7% text, 76.9% multimodal), followed closely by Gemini-3.1 Pro (91.4% and 78.7%), Gemini-3.6 Flash (91.0% and 74.8%) and GPT-5.6 Sol (89.9% and 76.7%). Accuracy significantly correlated with model size both for text-only and multimodal MCQs. Between model generations, the largest improvements in accuracy were seen for open-weight models whereas proprietary models showed only marginal gains. In error analysis, top-performing models exhibited highly concordant failure patterns, suggesting shared limitations on challenging cases. Frontier LLMs exhibit substantial specialist hematology knowledge across diverse subspecialist domains and clinical skill sets. Yet, despite high accuracy on board-style questions in hematology, continuous expert-on-the-loop output monitoring is paramount.

## Introduction

The introduction of ChatGPT^1^ has led to a Cambrian explosion of generative artificial intelligence (AI) systems permeating multiple knowledge-intensive domains, where they perform complex cognitive tasks that have traditionally required years of specialised human training. These transformer^2^-based large language models (LLMs) are trained on a vast amount of text and increasingly also incorporate multimodal data extending to images, video, and sound. Frontier developers of state-of-the-art (SOTA) commercial or open-weight chatbots currently engage in an arms race towards expert-level performance across an expanding number of benchmarks, evaluating factual knowledge retrieval, multi-step reasoning, mathematical and logical problem solving or software code generation.^3–8^ In the generalist AI domain, it has become a sport to design increasingly challenging benchmarks such as Humanity’s Last Exam^9^ or ARC-AGI-3^10^ to probe the limits of frontier models.

Their ability to encode medical knowledge^11^ along with problem-solving and multimodal reasoning have sparked a debate about the future role of LLMs in medicine. For instance, GPT-4o has been reported to significantly outperform medical students in the United States Medical Licensing Exam.^12^ Although LLMs increasingly saturate generalist medical benchmarks spanning student-level question answering (QA), literature and electronic health records summarisation as well as medical image classification and report generation (mostly on radiologic imaging)^13^, their evaluation on specialised medical knowledge remains insufficiently characterised. A recent survey by the American Medical Association has reported that 81% of clinicians use AI in their practice, more than doubling the rate reported in 2023^14^, yet appropriate regulatory frameworks for safe deployment of LLMs as medical devices are lacking^15^. This ‘shadow use’ is particularly concerning when LLMs are utilised to inform decision-making in high-stakes fields such as oncology and hematology. While the first studies evaluating LLMs in oncology emerge^16,17^, hematology – constituting more than 1.3 million new cases of hematologic malignancies per year globally with increasing incidence rates^18–20^ – remains uncharted terrain.

In this study, we comprehensively evaluate ten commercial and open-weight SOTA LLMs across five multimodal datasets, including 1,477 board-style questions in hematology spanning nine disease areas and six clinical skill sets.

## Methods

### Datasets

We evaluated 1,477 board-style multiple choice questions (MCQ) derived from five publicly accessible educational datasets curated from professional hematology societies, including the “Hematopoiesis Case Studies” from the American Society of Hematology (ASH)^21^, the educational resource platform of the British Society for Haematology (BSH)^22^ and the “Clinical Case of the Month” from the European Society for Blood and Marrow Transplantation (EBMT)^23^. An overview is provided in Fig. 1. BSH Multiple-Choice Questions (BSH-MCQ) and BSH Extended Matching Questions (BSH-EMQ) included 129 and 795 short text-only multiple-choice questions, each requiring assignment to a single correct answer option. BSH-Case Reports consisted of 486 multimodal (text and image data) long case vignettes. The ASH “Hematopoiesis Case Studies” dataset encompassed 40 text-only and four multimodal clinical cases. EBMT “Case of the Month” questions comprised 20 text-only and three multimodal case vignettes. Fig. S1 summarises the distribution of question types, question length and number of possible correct answers across these five datasets; Tab. S1 provides an overview of MCQ format per dataset. All reports are generated in accordance with TRIPOD-LLM^24^ guidelines.

**Fig. 1.**
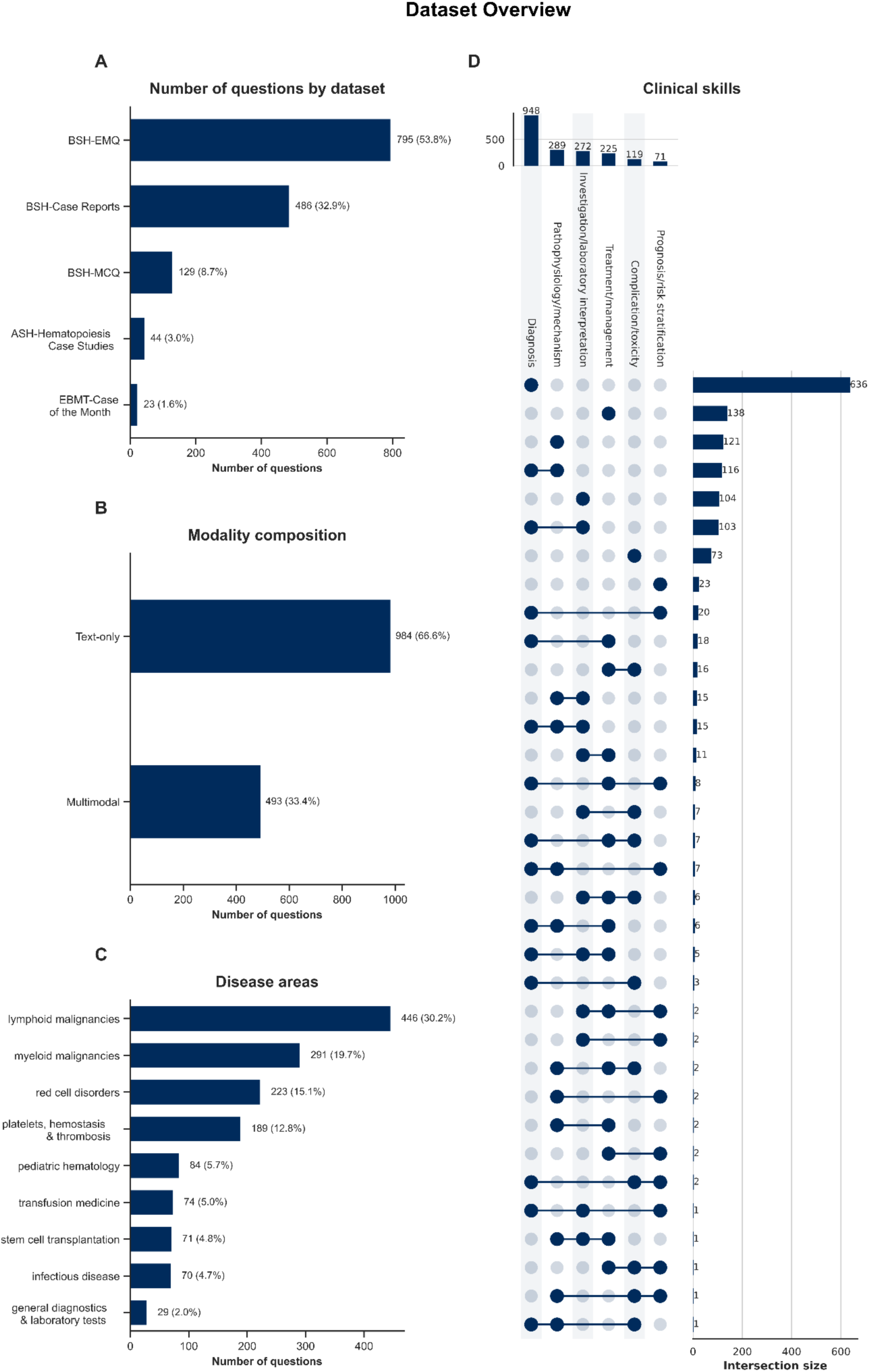
Benchmarking dataset composition. We collected 1,477 board-style exam questions from five sources (**A**), including both text-only as well as multimodal (text and images) case vignettes (**B**). These cases were further categorised into disease areas (**C**). In addition, cases were assigned up to three clinical skill sets required to solve the case (**D**).

To evaluate LLM performance across disease areas and clinical skills, we implemented a semi-automated pipeline for question categorisation (Fig. 1C, D). Specifically, we prompted a GPT-4.1-mini model to assign each question to one of several predefined hematological disease areas and one or more clinical skill categories. For each question, the model produced the three most likely categories along with their associated log-probabilities. These log-probabilities were converted to percentages and compared against a 90% confidence threshold. Categories that passed the threshold were automatically assigned to the dataset entries, while those below the confidence threshold were flagged for manual review by a hematologist (J-NE). Each question could receive up to three different clinical skill set categories, and therefore any entry failing to secure a confident top-1 match was expanded to include the next-highest ranks. Accordingly, each question was assigned to one of nine disease areas (lymphoid malignancies, myeloid malignancies, red cell disorders, platelet disorders, hemostasis and thrombosis, pediatric hematology, transfusion medicine, stem cell transplantation, infectious disease and general diagnostics and laboratory testing) as well as six clinical skill set categories (diagnosis, investigation and laboratory interpretation, treatment and management, pathophysiology and mechanism, complication and toxicity, and prognosis and risk stratification. Detailed category definitions are explained in Tab. S2 and S3.

### Large Language Models

We benchmarked ten SOTA LLMs, including four open-weight models and six proprietary models (Tab. 1). All models were served via Requesty (Requesty Ltd., London, United Kingdom) or OpenRouter (OpenRouter, Inc., New York City, United States of America) as API routing platforms, each directing requests to the respective model provider.

To ensure reproducibility, inference environments for all LLMs were standardised by constraining sampling parameters to minimise output randomness. The *temperature* was set to 0.0, to reduce sampling variability. The only exception was Kimi K3, which accepts only a temperature of 1.0. A *maximum token length* of 8192 was set to ensure comparable generation conditions across models, control inference cost, and prevent excessively long or degenerate completions from affecting downstream answer extraction and evaluation.

**Tab. 1.** Large Language Models used in the study. Included parameters are: Size in billions (*no official numbers available, community estimation), open or proprietary models, and knowledge cut-off date (**estimated, no official information available).

| LLM | Model Size (B) | Open-weight vs. Proprietary | Knowledge cut-off date | Last accessed on |
| --- | --- | --- | --- | --- |
| Anthropic Claude Opus 4.6 | ~5000 (~150 active)* | proprietary | May 2025 | August 7th 2026 |
| Anthropic Claude Opus 5 | ~5000 (~150 active)* | proprietary | May 2026 | August 7th 2026 |
| Google Gemini-3.1 Pro | ~3000* | proprietary | January 2025 | August 10th 2026 |
| Google Gemini-3.6 Flash | ~500* | proprietary | March 2026 | August 7th 2026 |
| Kimi K2.5 | 1000 (32 active) | open | April 2024 | August 10th 2026 |
| Kimi K3 | 2800 (104 active) | open | January 2026** | August 10th 2026 |
| OpenAI GPT-5.4 | ~3000 (~150 active)* | proprietary | August 2025 | August 10th 2026 |
| OpenAI GPT-5.6 Sol | ~5000 (~150 active)* | proprietary | February 2026 | August 7th 2026 |
| Z.ai GLM-4.6V | 106 | open | June 2025 | August 10th 2026 |
| Z.ai GLM-5V-Turbo | ~744 (~40 active)* | open | October 2025 | August 10th 2026 |

### Prompting and output evaluation

All LLMs were evaluated with zero-shot inference with a standardised prompt template, containing a system instruction providing the models with an experiment context and an expected answer format. Two different prompts were created depending on whether the dataset had questions with either one or more than one possible correct answers. When testing on MCQs with multiple possible correct answer options, the models were given the possibility to choose more than one correct answer, but were not required to do so. For the remaining MCQs, models were prompted to choose only one possible answer. After the system instruction, a dataset entry was presented with the case description and question (example in S1). The possible answers were prompted to the model in a randomised order to avoid positional token bias^25^. After inference, all raw model responses were post-processed and normalised to ensure consistent formatting for the downstream analysis pipeline.

The performance of all models was evaluated by measuring the accuracy of their responses per dataset and calculating the mean accuracy of all tested MCQs. Dataset entries with multiple correct answers were considered correct only if they were an exact match with the ground truth. In some cases, the models failed to provide a final output to the multiple-choice questions. These instances were considered as incorrect, reflecting the model’s inability to complete the given task.

We evaluated LLM responses by hematological disease area, clinical skill type, and modality, in order to identify patterns in incorrect model predictions. To quantify similarity in incorrect predictions between pairs of LLMs, we calculated the mean overlap of shared incorrect responses across all jointly misclassified questions. For single-possible-answer items, overlap was scored binarily (identical vs. non-identical incorrect response), whereas for multiple-possible-answer items, overlap was quantified using the Intersection over Union (IoU) of the incorrect response sets (detailed information in S2). We classified errors into five categories: incorrect answers in MCQs with single possibly correct answer (SPA), completely incorrect answers in MCQs with multiple possibly correct answers (MPA), over-selection, under-selection and mixed selection in questions with MPAs, and evaluated the distribution of error types across disease areas and the most common clinical skill types for all tested LLMs.

### Data availability

All datasets used for benchmarking are publicly available from BSH-MCQ (https://b-s-h.org.uk/education/bsh-education-resources/multiple-choice-questions), BSH-EMQ (https://b-s-h.org.uk/education/bsh-education-resources/extended-matching-questions), BSH-Case Reports (https://b-s-h.org.uk/education/bsh-education-resources/case-reports), ASH “Hematopoiesis Case Studies” (https://www.hematology.org/education/trainees/fellows/case-studies) and EBMT “Case of the Month” questions (https://www.ebmt.org/ebmt/news?combine=clinical+case).

## Results

### Generalist large language models solve board-style hematology multiple-choice questions with high accuracy

We evaluated ten SOTA LLMs using zero-shot prompting on 1,477 board-style MCQs across nine disease areas and six clinical skill sets. Overall, generalist proprietary models had the highest mean accuracy, yet many open-weight models followed closely behind. In text-only questions (n=984; 66.6% of total questions), Anthropic’s Claude Opus 5 ranked first with an accuracy of 92.7%, closely followed by Google’s Gemini-3.1 Pro and Gemini-3.6 Flash, OpenAI’s GPT-5.6 Sol, and Moonshot AI’s Kimi K3. Almost all models we evaluated – with the exception of Z.ai’s GLM-4.6V – passed the 80% correctness threshold (Fig. 2A). In multimodal (text plus images) questions, Gemini-3.1 Pro outperformed the other models with Claude Opus 5 and GPT-5.6 Sol ranking second and third. Importantly, multimodal case vignettes in our analyses stemmed predominantly from the “BSH-Case Reports” dataset (n=486; 98.6% of multimodal MCQs), which was not only multimodal but also allowed for multiple possibly correct answers. Hence, the performance drop in multimodal vs. text-based MCQs can likely be only partially attributed to multimodal reasoning failures as model accuracy was also affected by selecting the correct number of answer options instead of just one correct option in the text-only MCQs. Notably, newer generation models perform better than their older counterparts, with the exception of the Google models, as Gemini-3.6 Flash is a much smaller model than Gemini-3.1 Pro. Model size was significantly correlated with accuracy: Larger models significantly outperformed smaller models on text-only (Pearson r=0.77, *p*=0.009; Fig. 2B) and multimodal MCQs (Pearson r=0.79, *p*=0.007; Fig. 2C). Fig. S2 shows model accuracy by dataset.

**Fig. 2.**
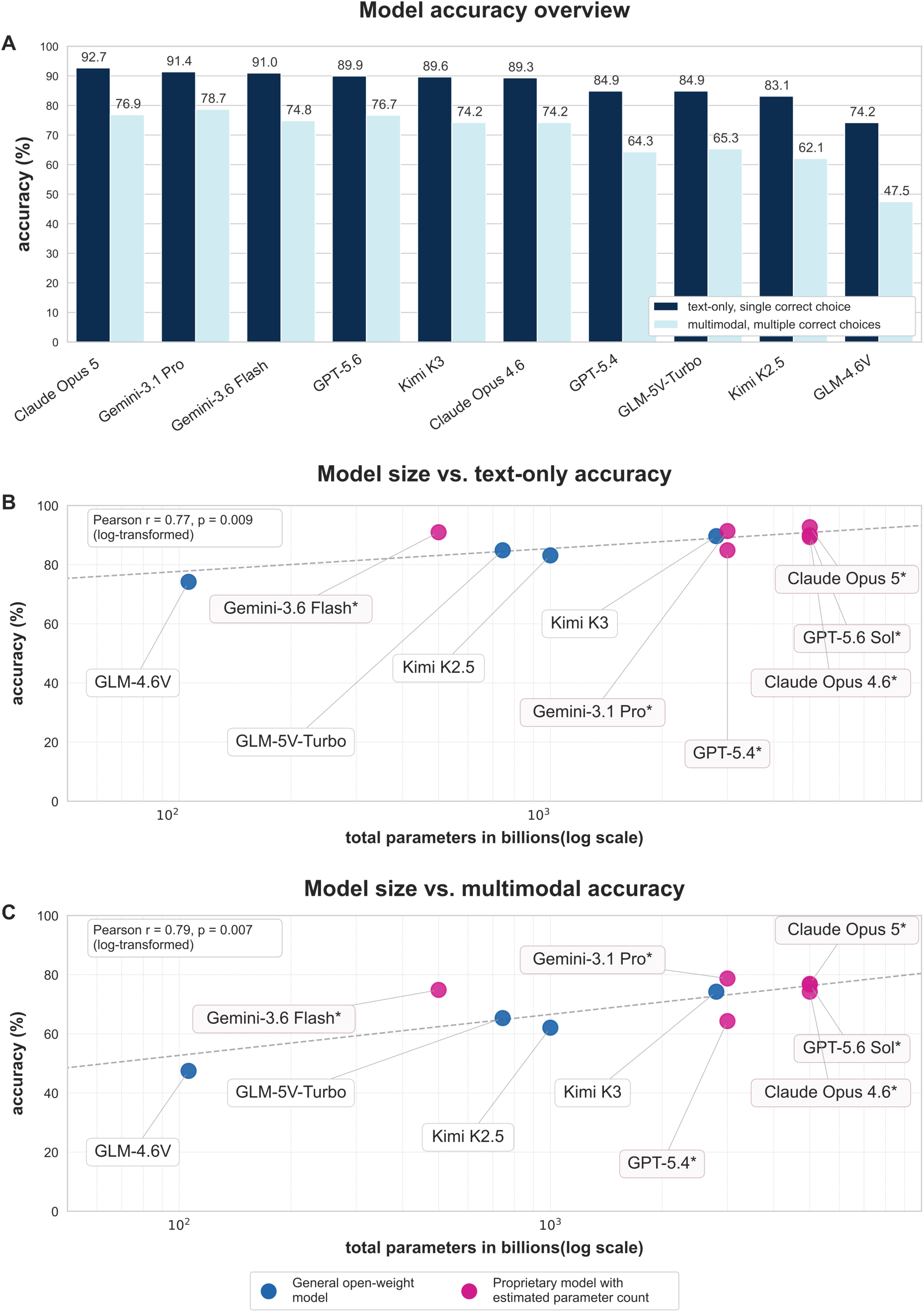
Proprietary models outperform open-weight models in hematological MCQs. Performance of ten state-of-the-art large language models (LLMs) was evaluated across 1,477 text-only and multimodal (text plus images) multiple choice questions (MCQs). (**A**). Proprietary models (Claude Opus 5, Gemini-3.1 Pro, GPT-5.6 Sol) slightly outperform open-weight models. Model size (in billion parameters) was correlated with accuracy, where on average larger models outperformed smaller ones for both text-only (**B**) and multimodal (**C**) MCQs. Of note, model size of the proprietary models is not officially disclosed by OpenAI, Anthropic and Google, but is based on community estimates (*).

### Model performance by disease category and clinical skill type

All MCQs were categorised into nine disease areas, pertaining to distinct hematological entities or subspecialists including pediatric hematology, transplantation, or transfusion medicine. We did not identify a distinct hematological entity or a subspecialty that individual models performed substantially better or worse in. In general, if a model had high overall accuracy, it also performed well in any given disease area without substantial outliers (Fig. 3, Fig. S3). With an average accuracy of 89.6% across all models, LLMs performed best on questions relating to transfusion medicine (n=74 MCQs), with the top four models (Claude Opus 5, Gemini-3.1 Pro, Gemini-3.6 Flash, GPT-5.6 Sol) reaching more than 95% accuracy. Questions in pediatric hematology followed with an average performance of 85.5% in n=84 MCQs. Models achieved similar accuracy of 83.9% and 82.1% in questions on platelets, hemostasis & thrombosis (n=189 MCQs) and in general diagnostics & laboratory tests (n=29), respectively, while achieving 81.8% accuracy in MCQs related to red cell disorders (n=223 MCQs). Marginal differences were also observed in areas of lymphoid malignancies (with 80.8% and n=446 MCQs) and infectious diseases (with 78.6% and n=70 MCQs). At the lower end of the spectrum, LLMs on average only achieved an accuracy of 77.9% and 76.9% on questions related to myeloid malignancies and stem cell transplantation, respectively. Across four of the five providers, the newer model generation outperformed their predecessor, with the highest gains of 10.9, 10.3 and 10.2 percentage points in MCQs related to red cell disorders, general diagnostics, and platelets, hemostasis & thrombosis, respectively. The notable exception was the Google model pair, in which Gemini-3.1 Pro outperformed the more recent Gemini-3.6 Flash with an average of 2.4 percentage points across all disease areas. The largest model improvement was observed in the GLM models, with the newer GLM-5V-Turbo outperforming GLM-4.6V with a mean 12.2 percentage points accuracy across all categories.

**Fig. 3.**
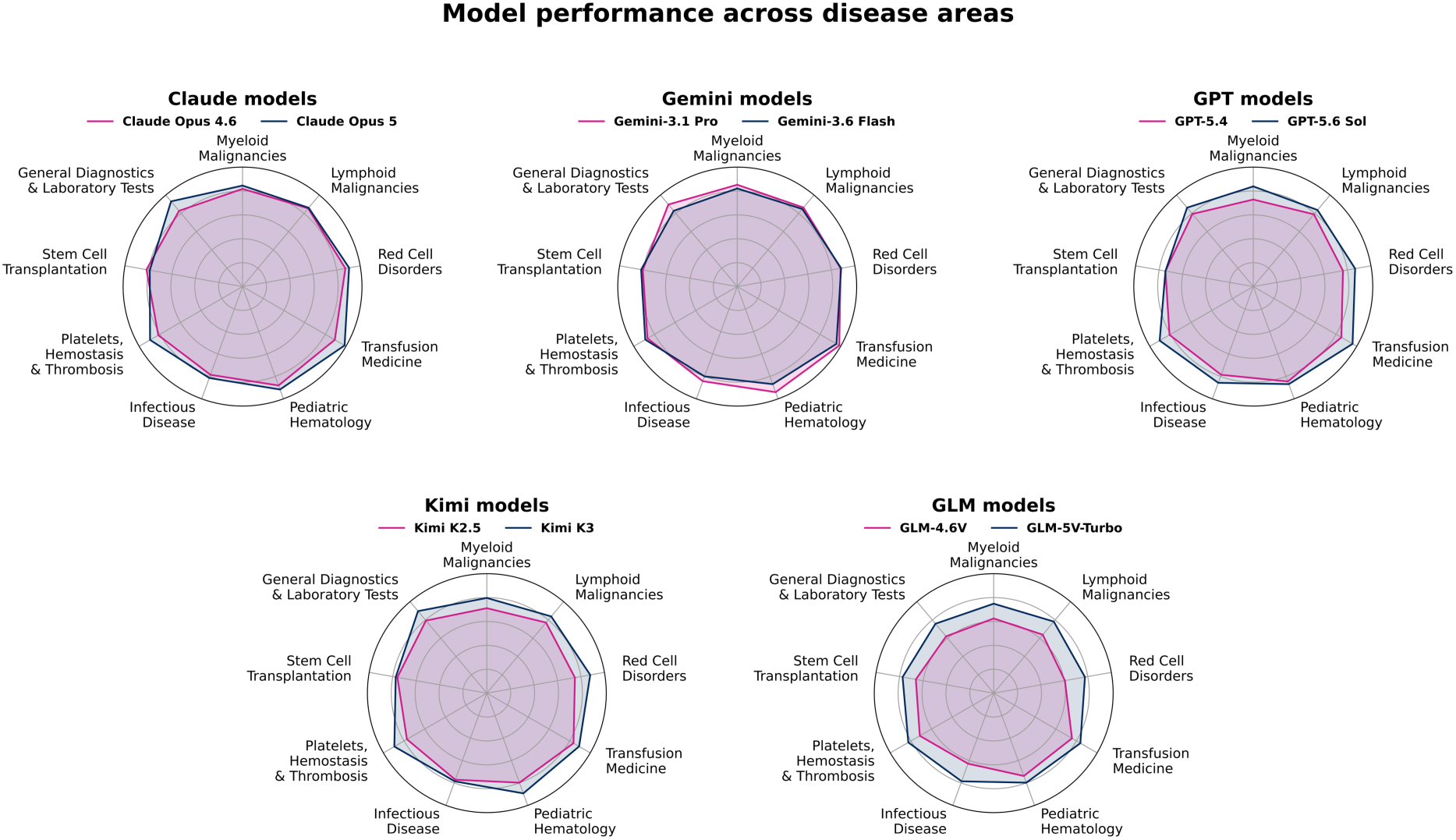
Model performance across nine disease areas. All MCQs were categorised according to disease area. Radar plots highlight disease area-specific accuracy for all ten models grouped by providers.

In addition to disease areas, all MCQs were also categorised according to clinical skills required to solve the case, where multiple skills could be assigned to a single MCQ. Again, models that performed well on average across all MCQs also performed well with regard to individual clinical skill sets (Fig. 4, Fig. S4). Most LLMs were able to correctly solve MCQs related to complication and toxicity management, with the five best performing models exceeding 85% correctness rate. Solving MCQs requiring diagnostic skills or interpretation of laboratory findings, only Gemini-3.1 Pro and Claude Opus 5 reached 86% correctness rate for both skill sets, while Gemini-3.6 Flash, GPT-5.6 Sol, Claude Opus 4.6 and Kimi K3 closely followed with 84-86% for diagnostics and 81-83% for interpreting laboratory findings. The same marginal differences were found for MCQs pertaining to treatment selection and therapeutic management, where these top performing models achieved 80-85% correctness rate. MCQs demanding an understanding of pathophysiology and underlying disease mechanisms were solved with over 81% correctness rates by Gemini-3.1 Pro, GPT-5.6 Sol, Claude Opus 5 and Gemini-3.6 Flash, closely followed by Claude Opus 4.6 (80.6%) and Kimi K3 (80.3%). Lastly, MCQs demanding risk stratification or prognostication were the hardest to solve, with two models falling in the 55-65% correctness range, while Claude Opus 5 and Gemini-3.1 Pro achieved 84.5% and 83.1%, respectively, and Gemini-3.6 Flash and Claude Opus 4.6 tied at 81.7%. Of note, the open-weight models evaluated here again showed the highest improvement between generations, with GLM-5V-Turbo improving on GLM-4.6V with an average of 13.5 percentage points across all clinical skills. The differences in proprietary models ranged on average between 1.8 and 5.5 percentage points.

**Fig. 4.**
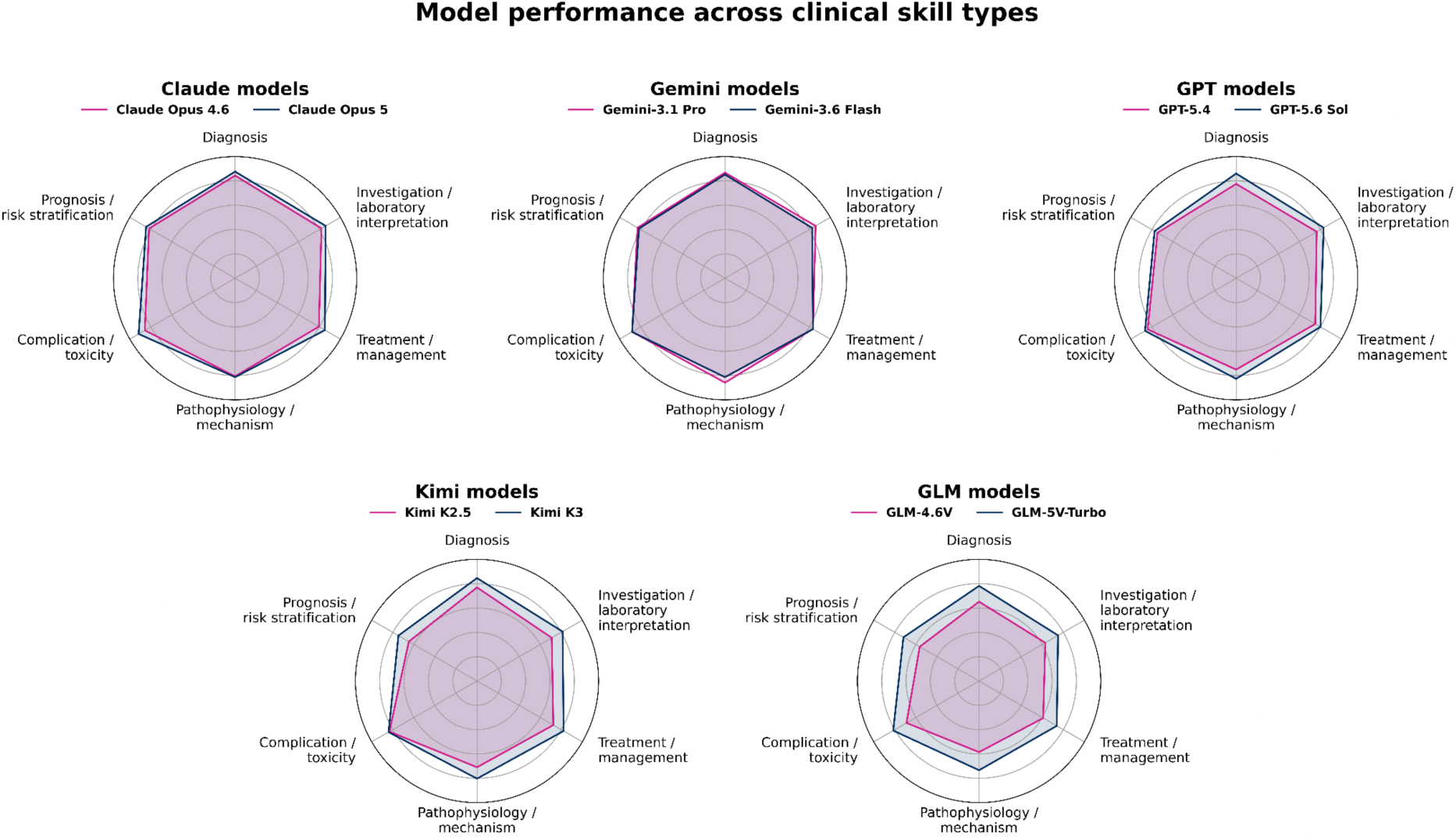
Model performance across clinical skill types. All MCQs were categorised according to at least one clinical skill. Radar plots highlight clinical skill-specific accuracy for all ten models grouped by providers.

### Top performing models make the same mistakes while low-performing models are heterogeneous in failure modes

Out of the ten models, both GPT models responded to every query, whereas both Kimi models had an average of 3.9% refusal rate. The other six models had a refusal rate of less than 1% (Fig. S5). Next, we investigated the overlap of incorrectly answered MCQs across all models, not including non-answered questions (Fig. S6). The top three performing models incorrectly answered between 177 and 204 MCQs with pairwise overlap in incorrectly answered MCQs ranging from 60.5% to 73.4%. For example, Claude Opus 5 incorrectly answered 185 MCQs, while Gemini-3.1 Pro incorrectly answered 177 MCQs. Claude Opus 5 shared 60.5% of its incorrectly answered MCQs with Gemini-3.1 Pro, while the latter shared 63.3% of its incorrectly answered MCQs with Claude Opus 5. The same pattern was observed for other top performing models, indicating that incorrectly answered MCQs were not primarily isolated failures of individual models, but instead corresponded to a common subset of challenging MCQs. The errors of more performant models were largely a subset of the errors of less performant models.

Additionally, we investigated the chosen incorrect answer options of the individual LLMs to assess a pattern in failure modes. The pairwise error similarity matrix in Fig. 5 (calculated as defined in S2) shows a structured distribution of model failures across the evaluation benchmark, where top models often selected the same incorrect answer options. In particular, the top performing models (Claude Opus 5, Gemini-3.1 Pro, Gemini-3.6 Flash and GPT-5.6 Sol) showed high error similarity, consistently selecting the same incorrect answer options, reaching a maximum overlap of 76.7% between Gemini-3.1 Pro and Gemini-3.6 Flash. Notably, when non-answered questions are not considered, Kimi K3’s failure rate drops from 229 to 163 incorrect MCQs, and demonstrates high overlap of answers with the top performing models, reaching 78.0% overlap of errors with Gemini-3.1 Pro. In contrast, lower-performing models exhibit more heterogeneous failure patterns, with weaker error similarity both relative to high-performing models and among themselves. This suggests that lower accuracy models not only fail across a broader set of examples, but also exhibit divergent errors, whereas higher accuracy models share common failure modes.

**Fig. 5.**
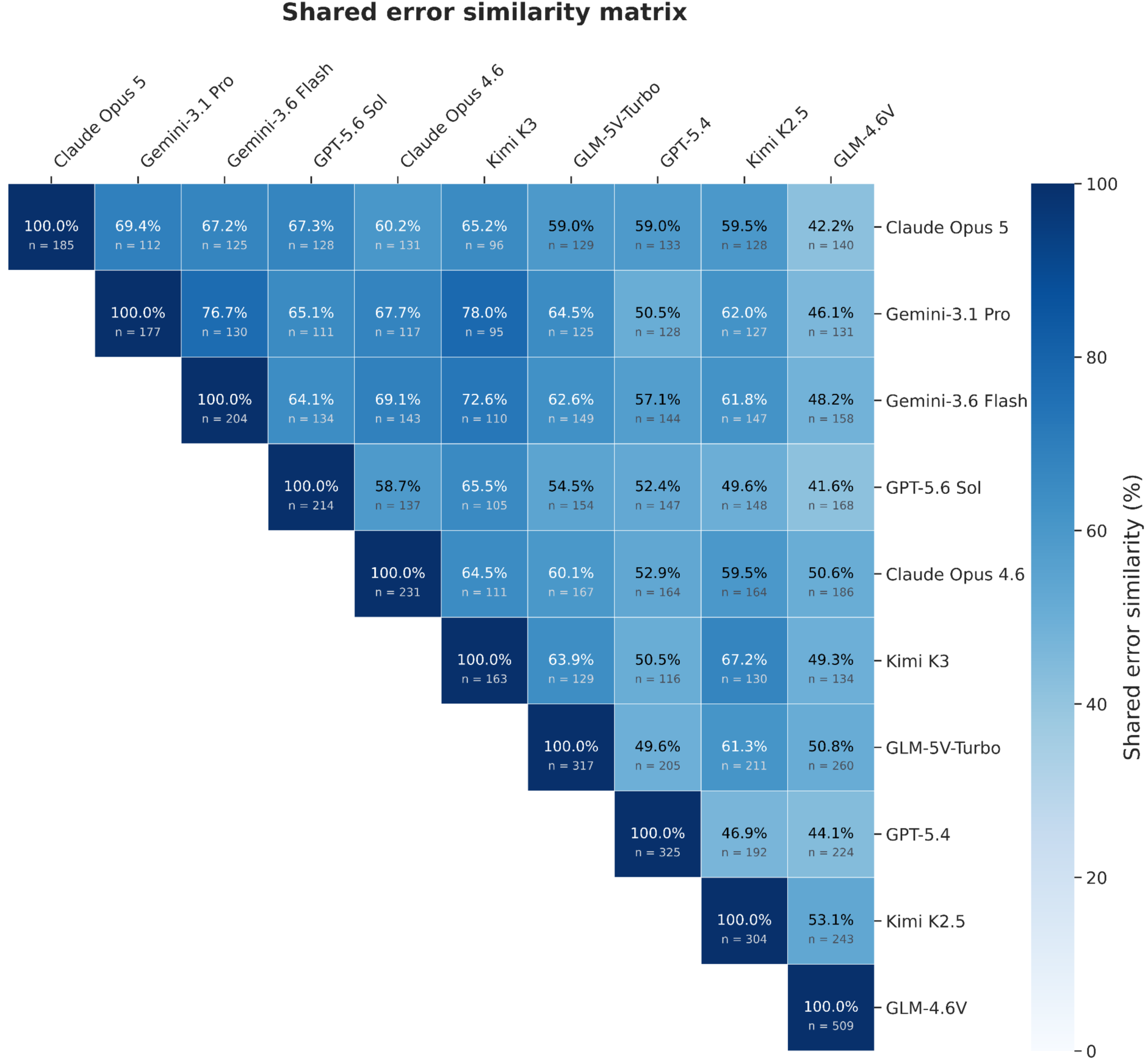
Shared error similarity matrix. Pairwise comparison of mistake alignment on questions where both models made an incorrect prediction. An Intersection over Union of errors is given as a percentage. In annotations, *n* indicates the number of shared MCQs with incorrect predictions. Sorted from highest to lowest mean accuracy.

Expanding on this analysis, we examined the distribution of error types in disease areas (Fig. 6 and Tab. S4) and clinical skill types (Fig. 7 and Tab. S5) across all ten models. In questions with multiple possibly correct answers, the most frequent error type was under-selection, i.e. models selected only a subset of the correct answer options, while omitting one or more correct options. Over-selection of answer options, i.e. selecting all correct answers plus incorrect answer options, was more frequent than mixed errors where models selected at least one correct and at least one incorrect answer option. Kimi K3 showed the highest output of completely incorrect answers in questions with multiple possible answers, spread evenly between disease areas. This finding partially reflects Kimi’s higher refusal rate, as refusals were classified as completely incorrect responses in our error taxonomy. For the other models, we did not find a pattern or tendency to over- or under-select answer options.

**Fig. 6.**
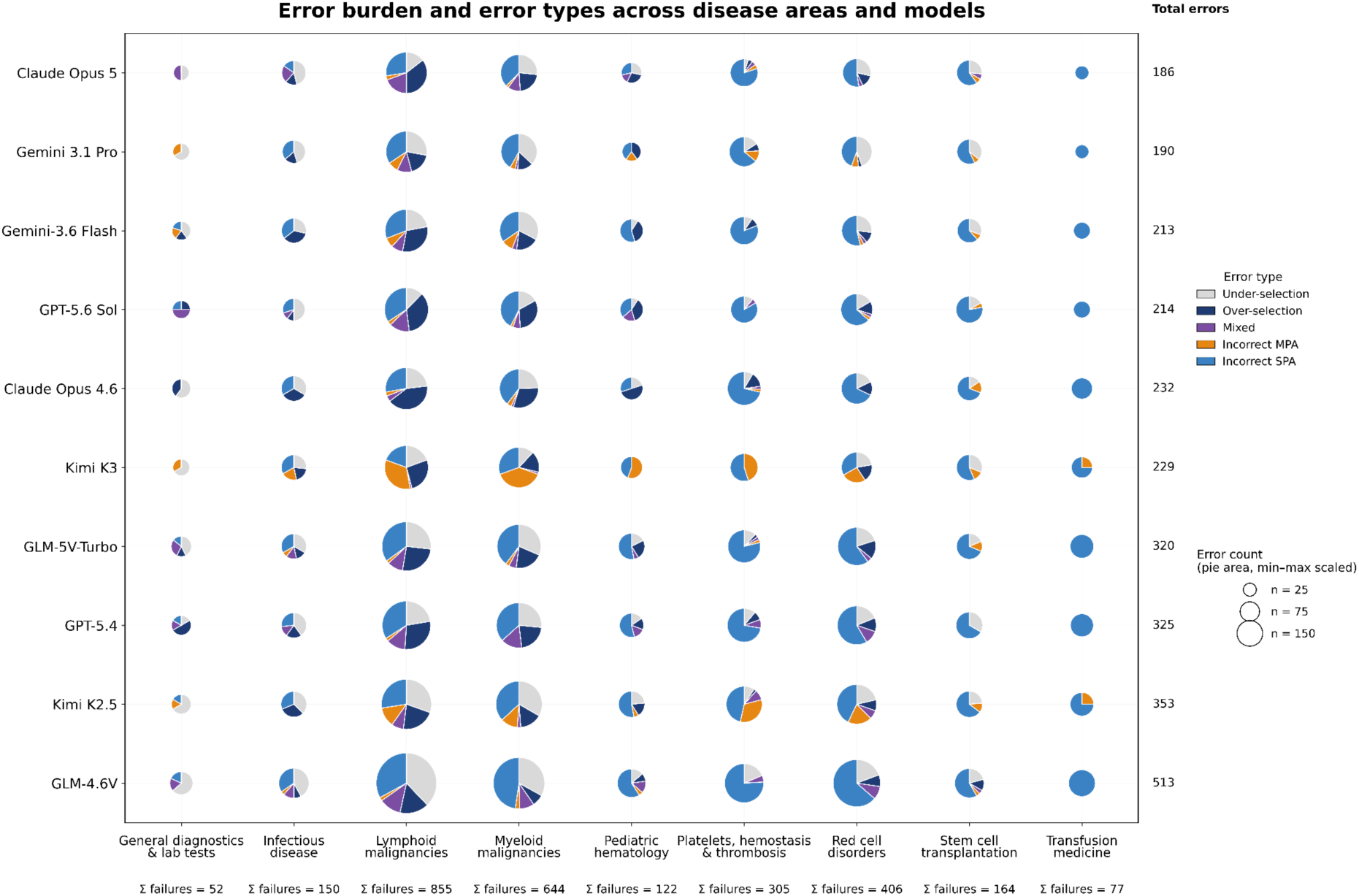
Error type distribution in nine disease areas. Model errors were classified as five error types: under-selection (model answer is a subset of the ground truth), over-selection (model answer is a superset of the ground truth), mixed selection (model answer contains partial set of the ground truth and partial incorrect answers), incorrect multiple possibly correct answers (MPA, model answer is completely incorrect in questions with multiple possible answers) and incorrect single possibly correct answer (SPA, model answer is completely incorrect in questions with single possible answer). Pie chart size shows the number of errors of each model in a given disease area. Total count of errors per model are listed on the right y-axis. Detailed information is given in Tab. S4.

For clinical skill types, MCQs related to toxicity were the only ones where models were prone to over-selection compared to the other categories where under-selection was the most frequent error type. Error types were more homogeneous between generations of the same model rather than between different models of the same generation.

**Fig. 7.**
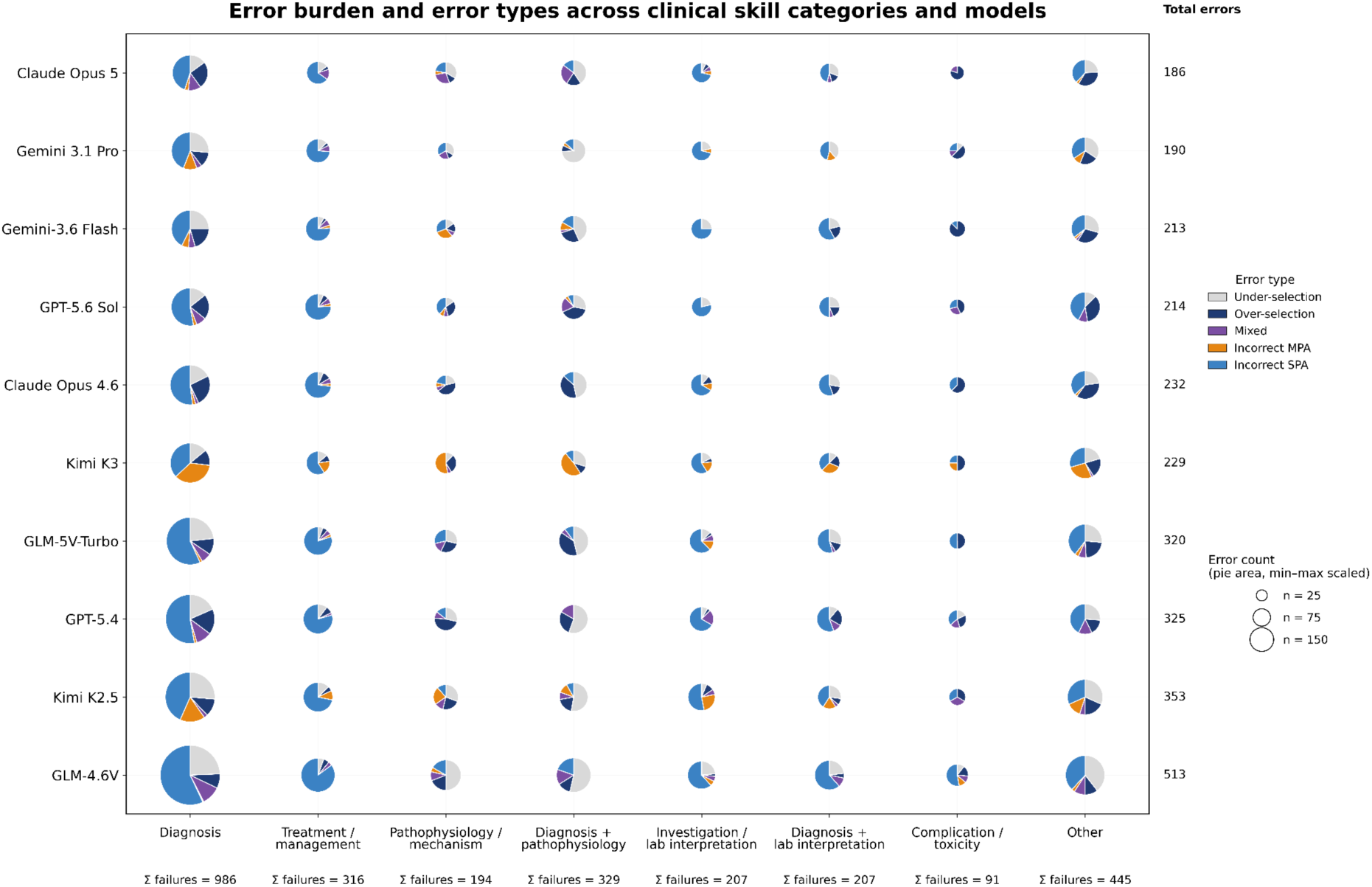
Error type distribution across clinical skill types. Model errors were classified as five error types: under-selection (model answer is a subset of the ground truth), over-selection (model answer is a superset of the ground truth), mixed selection (model answer contains partial set of the ground truth and partial incorrect answers), incorrect multiple possibly correct answers (MPA, model answer is completely incorrect in questions with multiple possible answers) and incorrect single possibly correct answer (SPA, model answer is completely incorrect in questions with single possible answer). Pie chart size shows the number of errors of each model in a given clinical skill. Total count of errors per model are listed on the right y-axis. Detailed information is given in Tab. S5.

## Discussion

We evaluated ten SOTA LLMs across 1,477 hematology board-style MCQs, assessing overall accuracy, as well as disease area-specific and clinical task-specific performance, and evaluated incorrect responses for failure modes. We found proprietary frontier models (Claude Opus 5, Gemini-3.1 Pro, Gemini-3.6 Flash and GPT-5.6 Sol) to perform exceptionally well in hematological MCQs across all disease areas, exhibiting broad clinical skills required to solve complex MCQs. Open-weight model performance fell only slightly behind the proprietary models. In general, larger models outperformed smaller models and – with the exception of the larger Gemini-3.1 Pro outperforming its smaller successor Gemini-3.6 Flash – newer models outperformed their predecessors. This performance jump between model generations was most pronounced in open-weight models while proprietary models had smaller performance jumps between generations. Interestingly, a newer generation open-weight model, Kimi K3, outperformed the previous generation Claude Opus 4.6 highlighting that open-weight models currently lag only months behind commercial models^26^. In error analysis, we did not find major differences for error modes between disease domains and clinical skills between all models, suggesting uniform performance across hematological subdomains and failure pertaining more to challenging cases rather than blind spots in specific domains or skill sets. Further, most top performing models made mistakes on the same MCQs and often selected the identical incorrect answer options, suggesting failure on a subset of especially challenging MCQs.

In oncology, a review of 34 studies^27^ revealed that LLMs are predominantly evaluated by analysing output correctness in QA, either in accordance with expert evaluation via Likert-scales or in reference to ground truth sources, often focusing on common malignant entities such as prostate, gynecological, oropharyngeal, or lung cancer. Notably, the lack of standardised reporting and frequent updates to frontier models contribute to substantial heterogeneity across both models and studies. In contrast to oncology, systematic benchmarking of LLMs in hematology remains limited with only few existing examples for specific disease areas. For instance, Swoboda et al.^28^ assessed GPT-4o, o3, Claude 4 Sonnet, and DeepSeek-V3 compared to a custom multi-agent system on 30 cases in myelodysplastic neoplasms (MDS), reporting major factual errors in 24% of cases for generalist LLMs. Moll et al.^29^ evaluated a retrieval-augmented generation (RAG) system based on a gpt-oss-120b backbone in multiple myeloma. Using a template with 48 standardised questions pertaining to treatment status, they queried 469 patient-question pairs from a single centre cohort with 100 patients, reporting a 79.6% concordance rate with expert annotations. Although discordance rates were similar to inter-expert discordance, LLM errors were associated with more severe safety implications.

Our study has several limitations. LLMs evaluated in our study were considered state of the art at the time of inference. However, the rapid pace at which frontier models are released and updated necessitates continuous benchmarking and community-based evaluation. Further, not all frontier models are uniformly accessible around the globe as, for instance, the US government compelled Anthropic to restrict access to Claude Fable 5 and Mythos 5^30^. Additionally, OpenEvidence AI, a popular medical model in the US, is currently unavailable in the EU and UK. With regard to multimodality, a caveat is that most multimodal MCQs also had multiple possible correct answer options, thereby obfuscating whether the performance drop in these MCQs came from subpar multimodal reasoning (compared to text-only) or from the arguably harder mode of MCQs. Next, our study focused on accuracy in QA, thereby simulating a hematology board exam. While most LLMs we evaluated demonstrated high accuracies, the question remains how well factual knowledge encoded in LLMs translates to clinical applicability. Although recent models such as Google’s AMIE or OpenAI’s o1-preview have been reported to outperform clinicians on diagnosis and medical reasoning^31,32^, evidence regarding real-world implementation and safety is still lacking. In a systematic review of 39 medical benchmarks, Gong et al.^33^ identified a persistent knowledge-practice gap, whereby older models excelling at standardised tests may still fail to translate that success into nuanced reasoning in real-world scenarios, thereby causing safety concerns. In support, Modi et al.^34^ reported that older models GPT-5, MedGemma-27B and OpenBioLLM-Llama3-70B exhibited substantial reasoning instability, reflected in broad accuracy shifts based on prompting strategy. As more clinicians incorporate LLMs into their practice^14^ and as agentic AI becomes integrated in clinical contexts^35^, awareness of their limitations and authoritative guidance on their appropriate usage is necessary. The recent ESMO guideline on the use of LLMs in Clinical Practice (ELCAP)^36^ provides a framework for standardising LLM use for healthcare providers, emphasising the need for critical evaluation rather than blind reliance on LLM outputs. Importantly, we demonstrated that model performance is not static, i.e. models that underperform in earlier iterations may improve substantially (in our analysis most pronounced for GLM models), whereas previously reliable models still warrant constant assessment (in our analysis most prominent with Google models) as updates may introduce new model behaviour. Analogous to post-market pharmacovigilance, hematology and oncology should devise frameworks for standardised, ongoing ‘LLM vigilance’ as these tools become increasingly integrated into clinical practice.

In summary, we evaluated ten SOTA LLMs across 1,477 hematology board-style MCQs, assessing overall accuracy as well as disease area-specific and clinical task-specific performance and evaluated model failure modes. We found frontier models to perform exceptionally well, exhibiting complex clinical reasoning skills on expert-level hematology MCQs. However, high scores on benchmark tasks do not guarantee safe implementation in a real-world context, warranting constant output monitoring by experts, regulatory oversight and authoritative guidance by professional societies.

## Data Availability

All datasets used for benchmarking are publicly available from BSH-MCQ (https://b-s-h.org.uk/education/bsh-education-resources/multiple-choice-questions), BSH-EMQ (https://b-s-h.org.uk/education/bsh-education-resources/extended-matching-questions), BSH Case Reports (https://b-s-h.org.uk/education/bsh-education-resources/case-reports), ASH Hematopoiesis Case Studies (https://www.hematology.org/education/trainees/fellows/case-studies), and EBMT Case of the Month (https://www.ebmt.org/ebmt/news?combine=clinical+case).

https://b-s-h.org.uk/education/bsh-education-resources/extended-matching-questions

https://b-s-h.org.uk/education/bsh-education-resources/case-reports

https://www.hematology.org/education/trainees/fellows/case-studies

https://www.ebmt.org/ebmt/news?combine=clinical+case

## Acknowledgements

The authors gratefully acknowledge the computing time made available to them on the high-performance computer at the NHR Center of TU Dresden. This centre is jointly supported by the Federal Ministry of Research, Technology and Space of Germany and the state governments participating in the NHR (www.nhr-verein.de/unsere-partner). This study was supported by a grant from the Deutsche Krebshilfe (German Cancer Aid, Project Number 70117128) to J-NE.

## Authorship Contributions

MR, MoB and J-NE conceptualised the study. MR collected multiple choice questions. MR and MoB implemented the benchmarking software pipeline. All authors evaluated model performance. MR, MoB and J-NE wrote the initial draft. All authors reviewed and edited the draft and agreed on the final version and the decision to submit for publication. All authors had access to all data and results and agreed to be accountable for all aspects of the study.

## Supplementary Methods

### S1 Prompt example

Example of a multimodal question with multiple possible answers from the EBMT-Case of the Month dataset with a prediction from Claude Opus 5 (and a post-processed answer):

### Prompt

You are an expert, board-certified hematologist-oncologist with extensive clinical and academic experience.You will receive a case description with a question and multiple-choice answers.Output ONLY the final choice as an integer number (1, 2, 3…). If multiple answers are correct, output them as a list of integers (eg [1, 3]). Format EXACTLY as: ‘Answer: <number>’ or ‘Answer: [<number>, <number>]’.

An 82-year-old woman with grade 3A, stage IV follicular lymphoma was treated with CD19 directed CAR T-cells (lisocabtagene maraleucel) after three prior lines of therapy. Baseline brain MRI was normal. The early post-infusion course was marked by grade 1 ICANS on day +6 and by the persistance of complete B-cell aplasia (0 × 10⁹/L) from day +7 and during all the follow-up, consistent with the expected on-target/off-tumor effect of CD19 directed CAR T-cells About 30 months after CAR T-cell therapy, she progressively developed cognitive symptoms, including dyslexia, dysgraphia, aphasia, and memory impairment. Brain MRI revealed asymmetric bilateral parieto-occipital T2-FLAIR hyperintensities. JC virus PCR on cerebrospinal fluid was positive at 12,844 IU/mL (4.11 log₁₀), confirming the diagnosis of progressive multifocal leukoencephalopathy.

At diagnosis, CAR T-cells were still detectable by qPCR (470 copies/10⁶ leukocytes; 2.67 log₁₀). Blood immunophenotyping showed persistent B-cell aplasia, hypogammaglobulinemia (4.0 g/L), and CD4⁺ T-cell lymphopenia (0.3 × 10⁹/L) with a marked reduction in naïve CD4⁺ T-cells (2%). Immune checkpoint blockade with nivolumab was initiated, combined with intravenous immunoglobulin. Treatment was well tolerated. After 3 cycles, JC viral load markedly decreased to a detectable but non-quantifiable level. MRI showed a dissociated response, with regression of all previous white matter lesions but progression of a left frontal lesion >1 cm (Figure 1). End treatment blood immunophenotyping showed persistent B-cell aplasia (0 × 10⁹/L) and CD4⁺ T-cell lymphopenia (0.38 × 10⁹/L) (Figure 2). One year after PML diagnosis and four years after CAR T-cell infusion, the patient showed significant cognitive function improvement, and remained alive with limited functional impairement.

‘url’: ‘https://www.ebmt.org/sites/default/files/inline-images/202611_Clinical%20Case_Figure1.jpg’, ‘detail’: ‘high’

`CAPTION: Brain MRI evolution under nivolumab treatment. Axial FLAIR images at baseline (A), after two cycles of nivolumab (B), and six months after treatment completion (C)`.

‘url’: ‘https://www.ebmt.org/sites/default/files/inline-images/202611_Clinical%20Case_Figure2.jpg’, ‘detail’: ‘high’

`CAPTION: Evolution of peripheral lymphocyte subsets during hematologic follow-up and after nivolumab`.

`Which immune abnormalities may have favored the development of progressive multifocal leukoencephalopathy in this patient?`

`Options`:

`5. Neutrophilia`
`4. Expansion of regulatory T cells`
`3. Hypergammaglobulinemia`
`2. Persistent B-cell aplasia`
`1. CD4⁺ T-cell lymphopenia`

**<u>`LLM Response`:</u>**

`Answer: [1, 2]`

**<u>`Post-processed Answer`:</u>**

`[1, 2]`

**<u>`Ground Truth`:</u>**

`[1, 2]`

### S2 Overlap of incorrect predictions between models

For an accurate representation of the similarity in incorrect model predictions, we calculated the Intersection over Union (IoU) of all incorrect responses made by all model pairs and converted their score to a percentage. This global IoU of two models can be defined as:

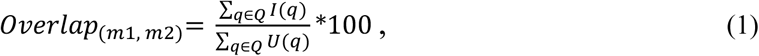

where *m*1 and *m*2 are two of the LLMs, *Q* is the set of questions, which both *m*1 and *m*2 answered incorrectly, I(q) is the intersection of errors for a given question q and U(q) is the union of errors for a given question q. An incorrect response is defined as a missing correct answer or a wrong answer given by a model. Examples are given for calculating an overlap between Gemini-3.1 Pro and GPT-

5.4 for two sample questions from the BSH-Case Reports:

<u>Example 1</u>: Ground Truth: {0, 2, 4}

Gemini-3.1 Pro Prediction: {0, 4} | GPT-5.4 Prediction: {0, 1, 4}

The error set for Gemini-3.1 Pro is {2} and for GPT-5.4 is {1,2}. Therefore *I*(*q*)_1_=1 and *U*(*q*)_1_=2.

<u>Example 2</u>; Ground Truth: {0, 1, 2, 4}

Gemini-3.1 Pro Prediction: {0, 3, 4} | GPT-5.4 Prediction: {1, 2, 3, 4}

The error set for Gemini-3.1 Pro is {1,2,3} and for GPT-5.4 is {0,3}. Therefore *I*(*q*)_2_=1 and *U*(*q*)_2_=4.

The overlap between Gemini-3.1 Pro and GPT-5.4 is then calculated as 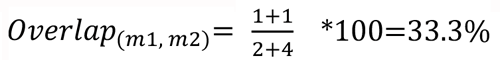.

## Supplementary Tables

**Tab. S1.**
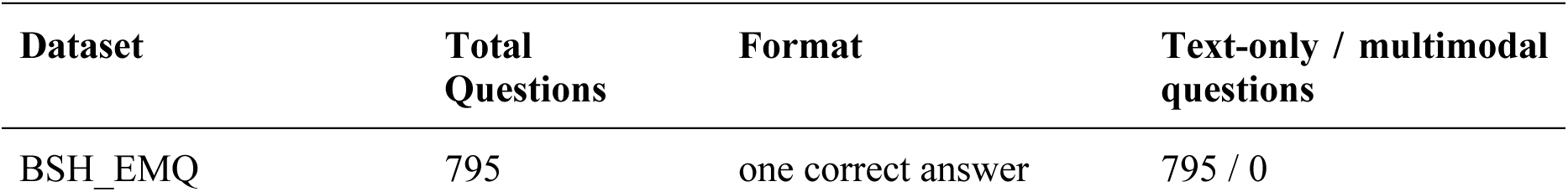
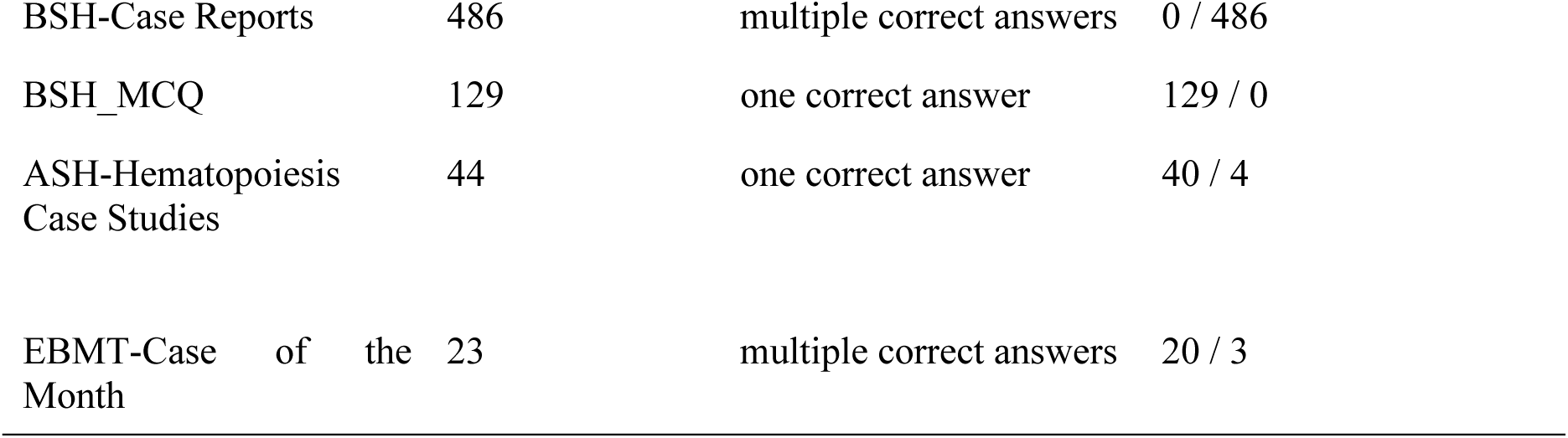
Datasets overview.

**Tab. S2.** Detailed definitions of disease area categories.

| Disease area | Definition |
| --- | --- |
| Myeloid Malignancies | AML, MDS and myeloproliferative neoplasms (CML, polycythaemia vera, essential thrombocythaemia, myelofibrosis). |
| Lymphoid Malignancies | Non-Hodgkin lymphoma, Hodgkin lymphoma, CLL, and plasma cell disorders (myeloma, MGUS) |
| Red Cell Disorders | Anemias; haemoglobinopathies; erythrocytosis. |
| Platelets, Hemostasis & Thrombosis | Platelet disorders (TTP, HIT, ITP), coagulation factor deficiencies, thrombophilia and VTE. |
| Pediatric Hematology | Pediatric-specific presentations, congenital disorders; neonatal hematology. |
| Transfusion Medicine | Blood components (red cells, platelets, FFP); transfusion reactions; compatibility testing. |
| Stem Cell Transplantation | Allogeneic and autologous HSCT; indications, conditioning, and complications. |
| Infectious Disease | Infections relevant to hematology: febrile neutropenia, fungal disease, malaria, HIV. |
| General Diagnostics & Laboratory Tests | Principles and lab techniques (morphology, bone marrow, molecular tests), general disease. |

**Tab. S3.** Detailed definitions of clinical skill types.

| Clinical skill | Definition |
| --- | --- |
| Diagnosis | Identifying the most likely diagnosis from history, examination, and/or investigation. |
| Investigation/laboratory interpretation | Selecting and interpreting diagnostic tests, such as flow cytometry, cytogenetics, and molecular assays. |
| Treatment/management | Selecting and ordering treatments, including medications, transplantation, and supportive care. |
| Pathophysiology/mechanism | Understanding the molecular or cellular basis of disease or drug action. |
| Complication/toxicity | Identifying or handling treatment side effects or disease-related complications. |
| Prognosis/risk stratification | Utilisation of prognostic scoring systems or risk stratification parameters to guide and optimise clinical decision-making. |

**Tab. S4.**
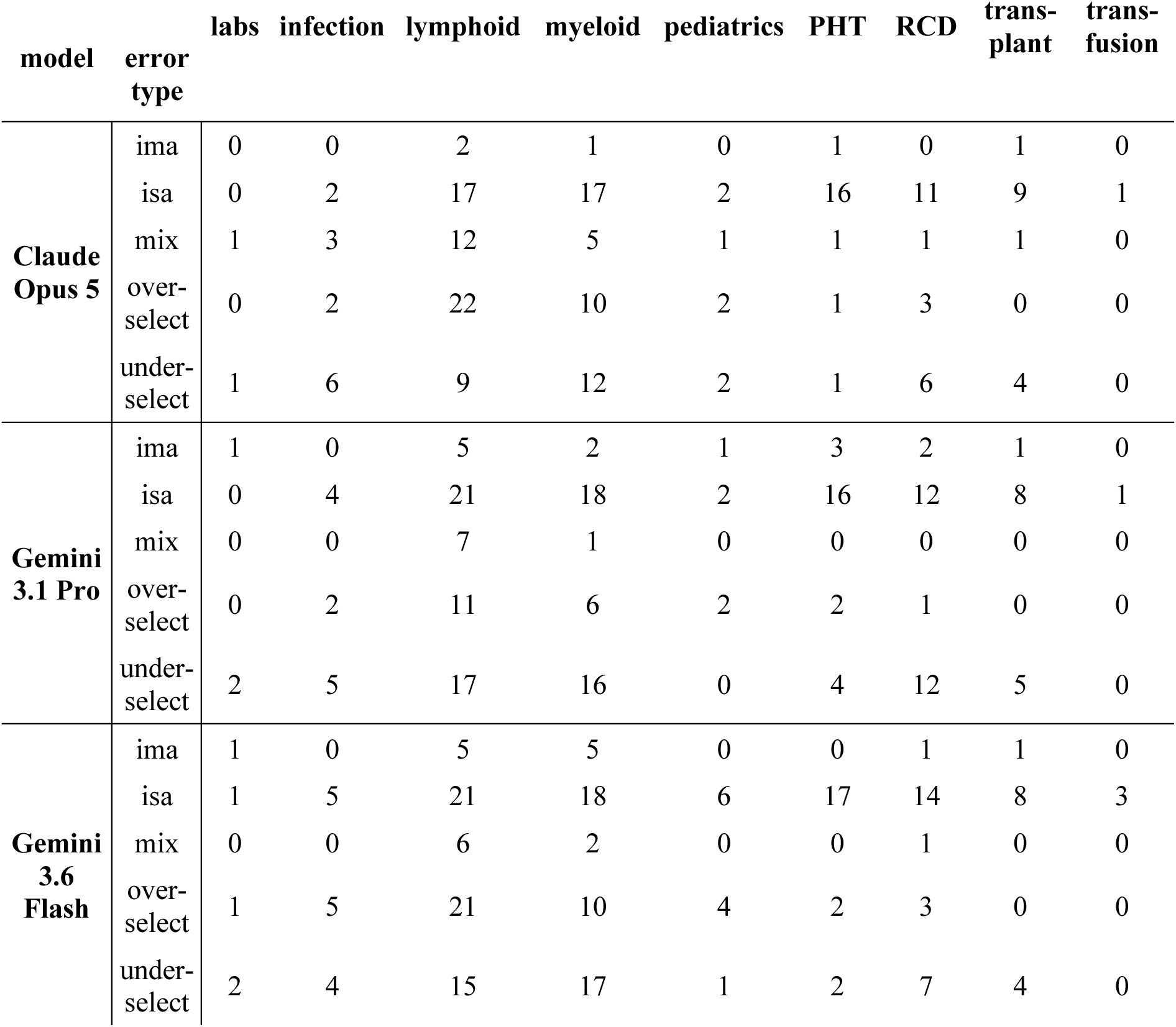
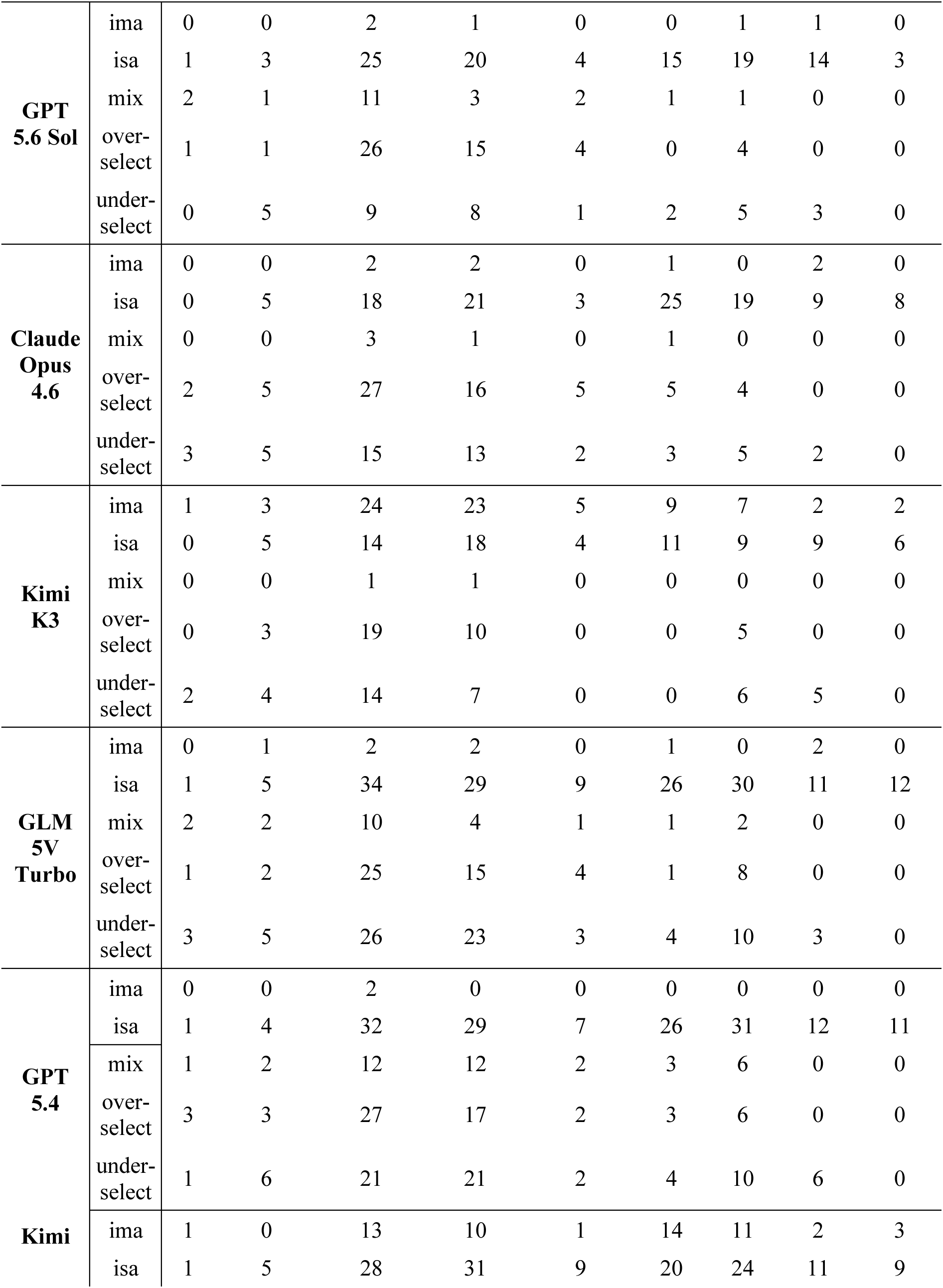
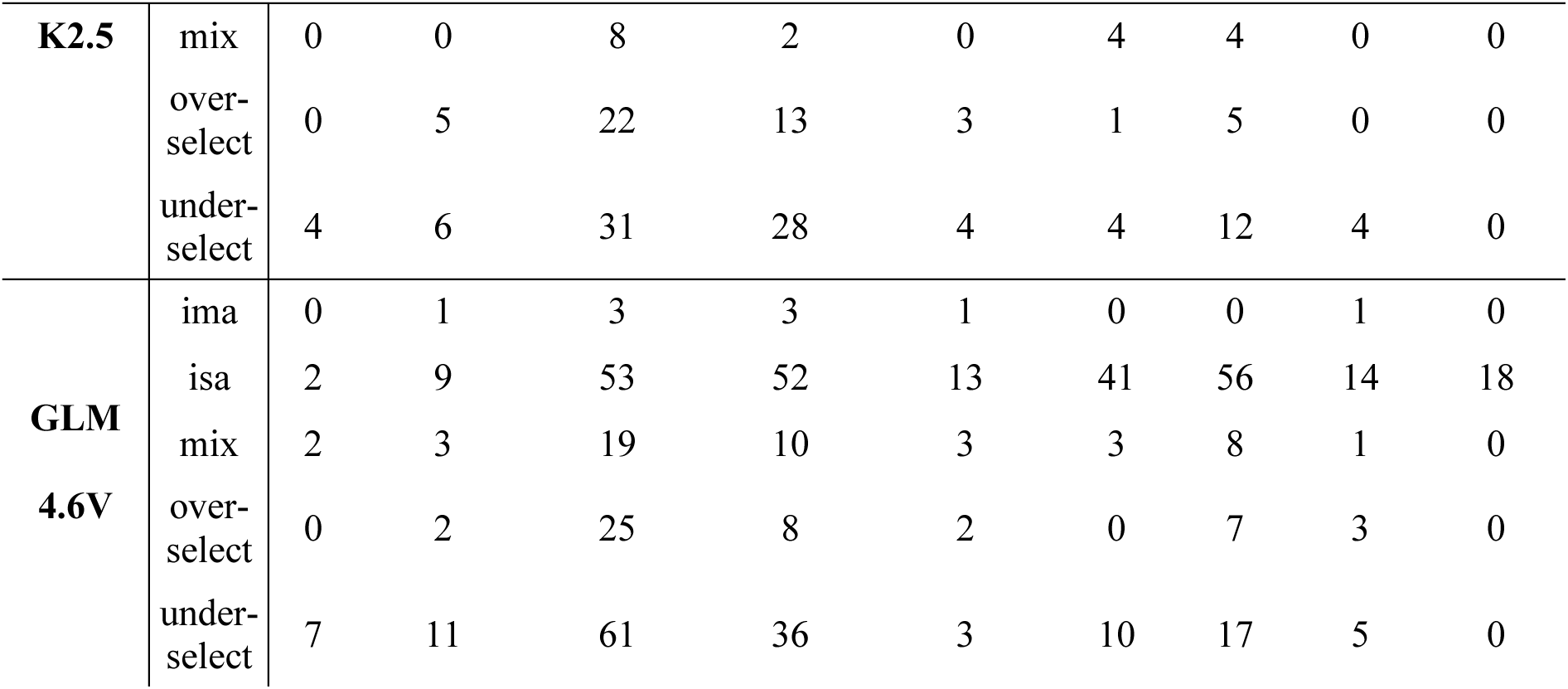
Detailed error types in disease areas. Disease areas: labs. = General Diagnostics & Laboratory Tests; **infection** = Infectious Disease, **lymphoid** = Lymphoid Malignancies, **myeloid** = Myeloid Malignancies, **pediatrics** = Pediatric Hematology, **PHT** = Platelets, Hemostasis & Thrombosis, **RCD** = Red Cell Disorders, **transplant** = Stem Cell Transplantation, **transfusion** = Transfusion Medicine. **Error types: ima** = incorrect multi-answer (disjoint prediction for multi-item ground truth); **isa** = incorrect single-answer (disjoint prediction for single-item ground truth); **mix** = mixed selection (partial overlap with ground truth); **over-select** = over-selection (superset containing all target and extra items); **under-select**, under-selection (strict subset omitting target items).

**Tab. S5.**
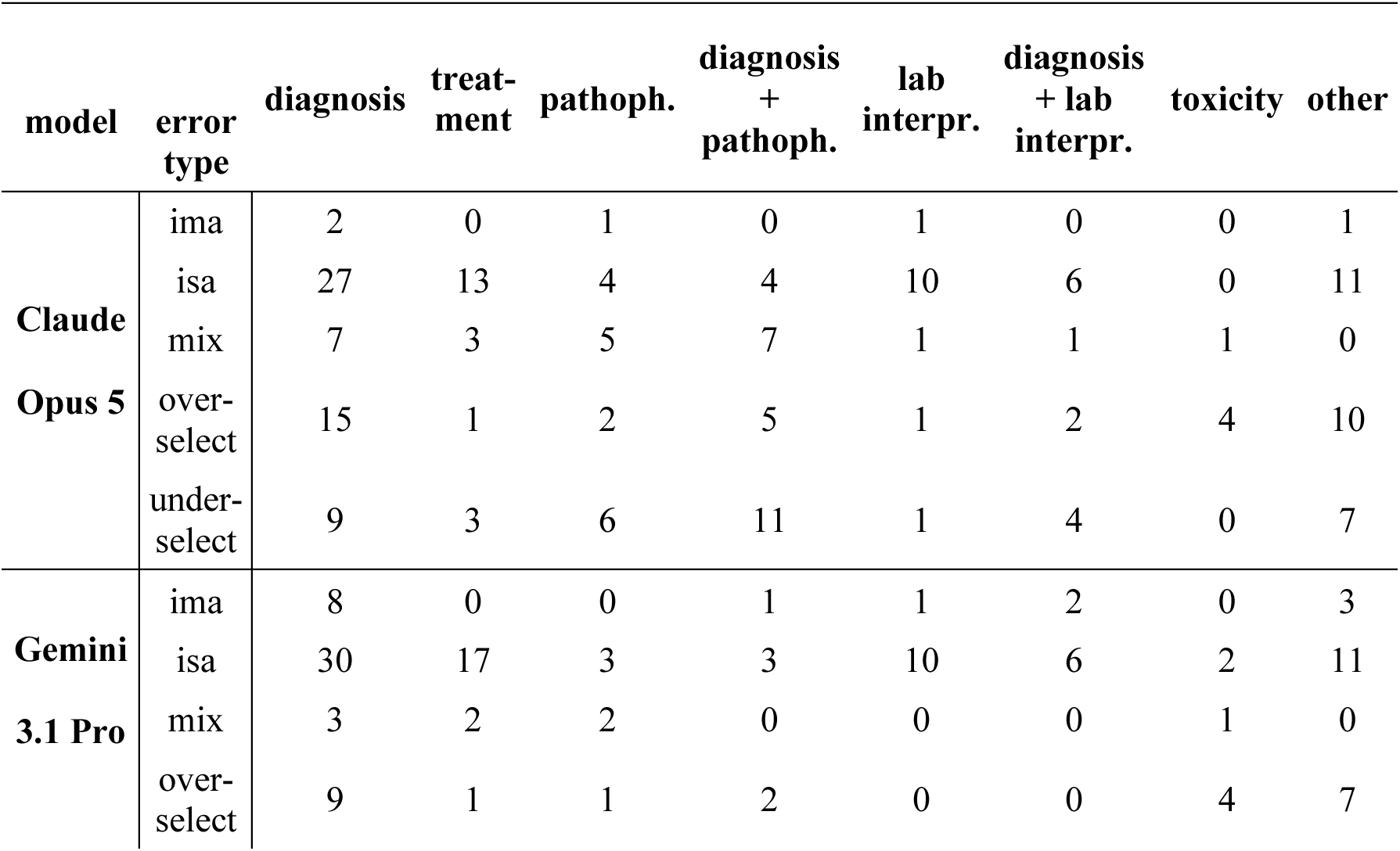
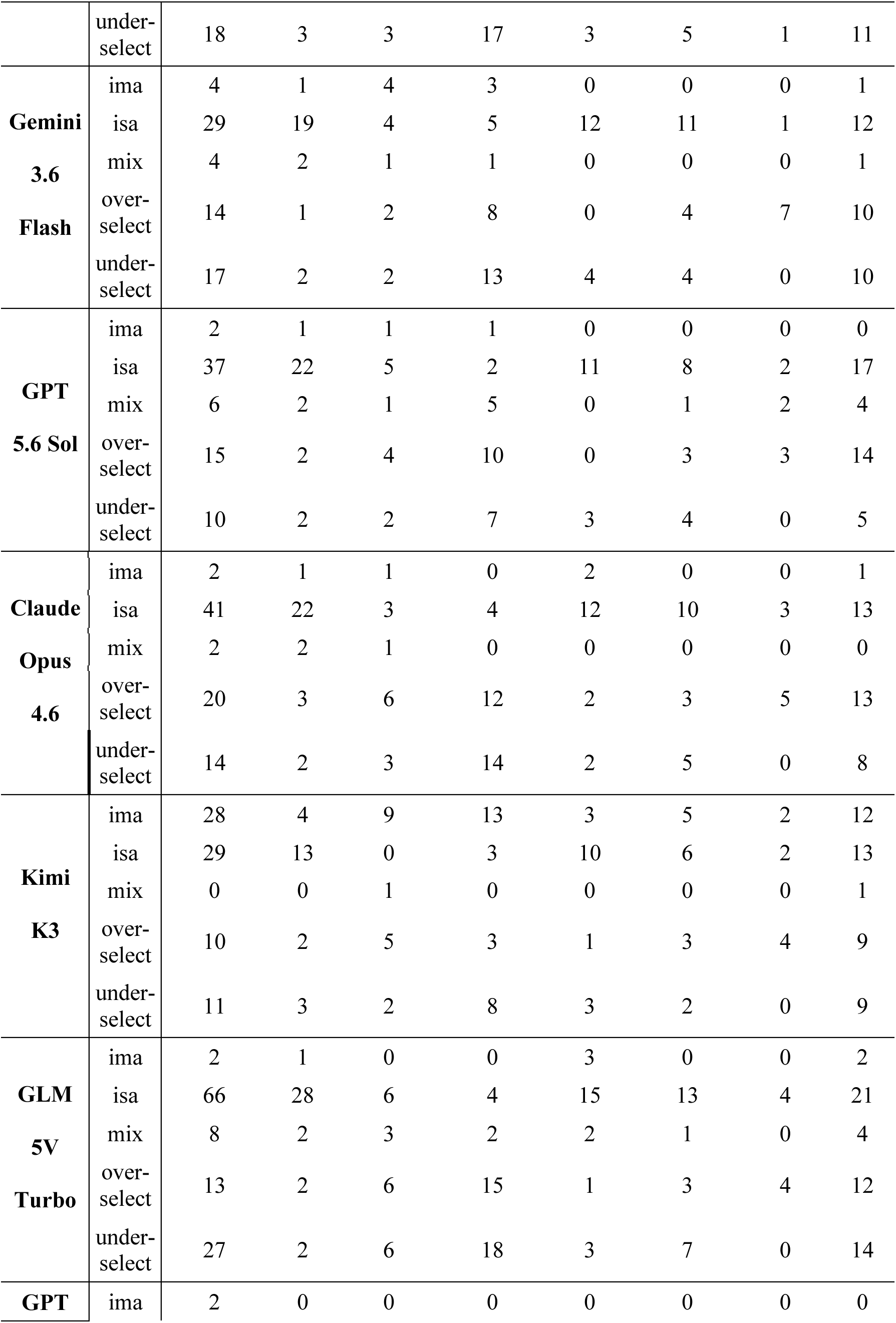
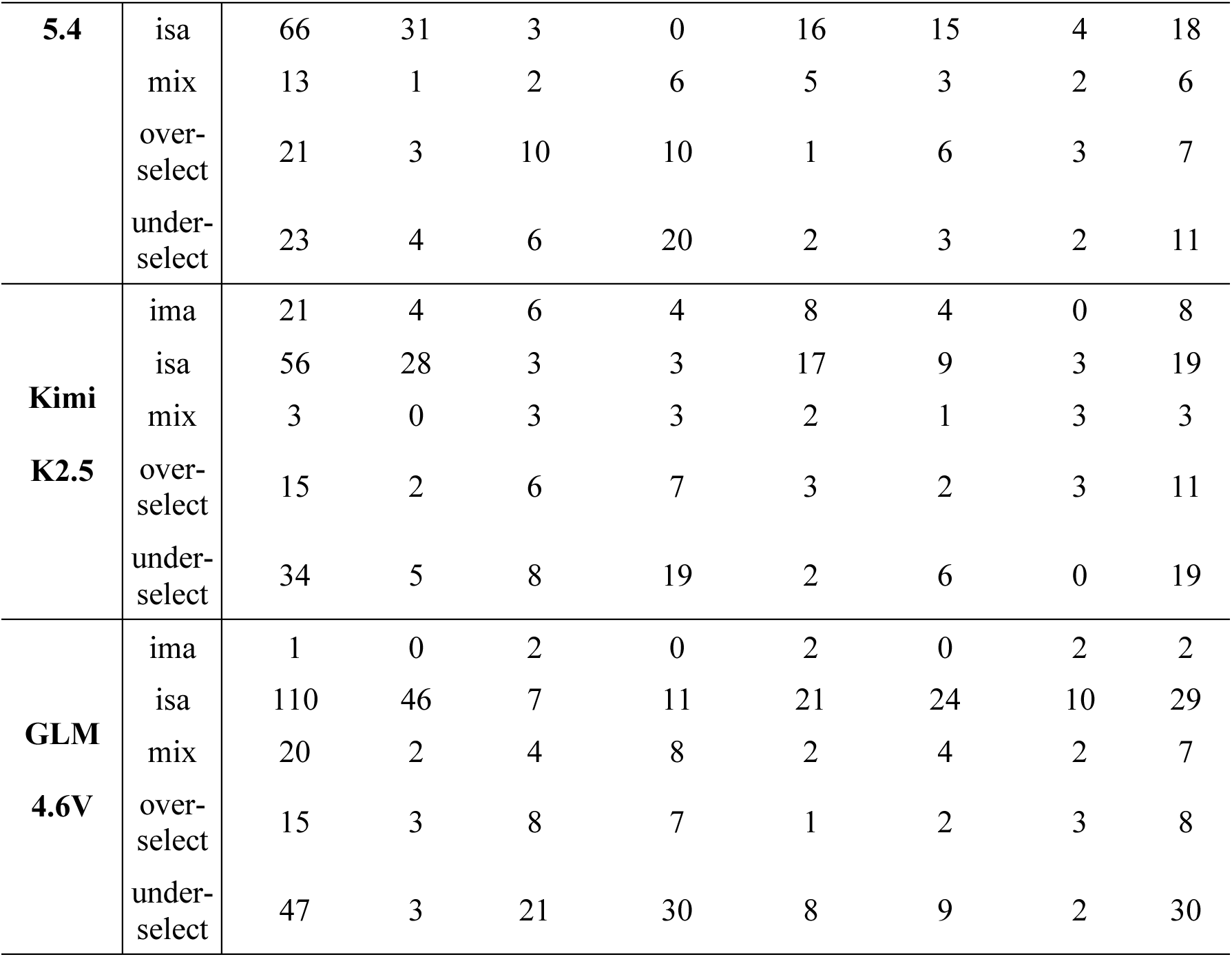
Detailed error types in clinical skill types. Clinical skills: diagnosis. = Diagnosis; **treatment** = Treatment/management; **pathoph.** = Pathophysiology/mechanism; **diagnosis + pathoph.** = Diagnosis & Pathophysiology/mechanism; **lab interpr.** = Investigation/laboratory interpretation; **diagnosis + lab interpr.** = Diagnosis & Investigation/laboratory interpretation; **toxicity** = Complication/toxicity; **other** = all other clinical skill types. **Error types: ima** = incorrect multi-answer (disjoint prediction for multi-item ground truth); **isa** = incorrect single-answer (disjoint prediction for single-item ground truth); **mix** = mixed selection (partial overlap with ground truth); **over-select** = over-selection (superset containing all target and extra items); **under-select**, under-selection (strict subset omitting target items).

## Supplementary Figures

**Fig. S1.**
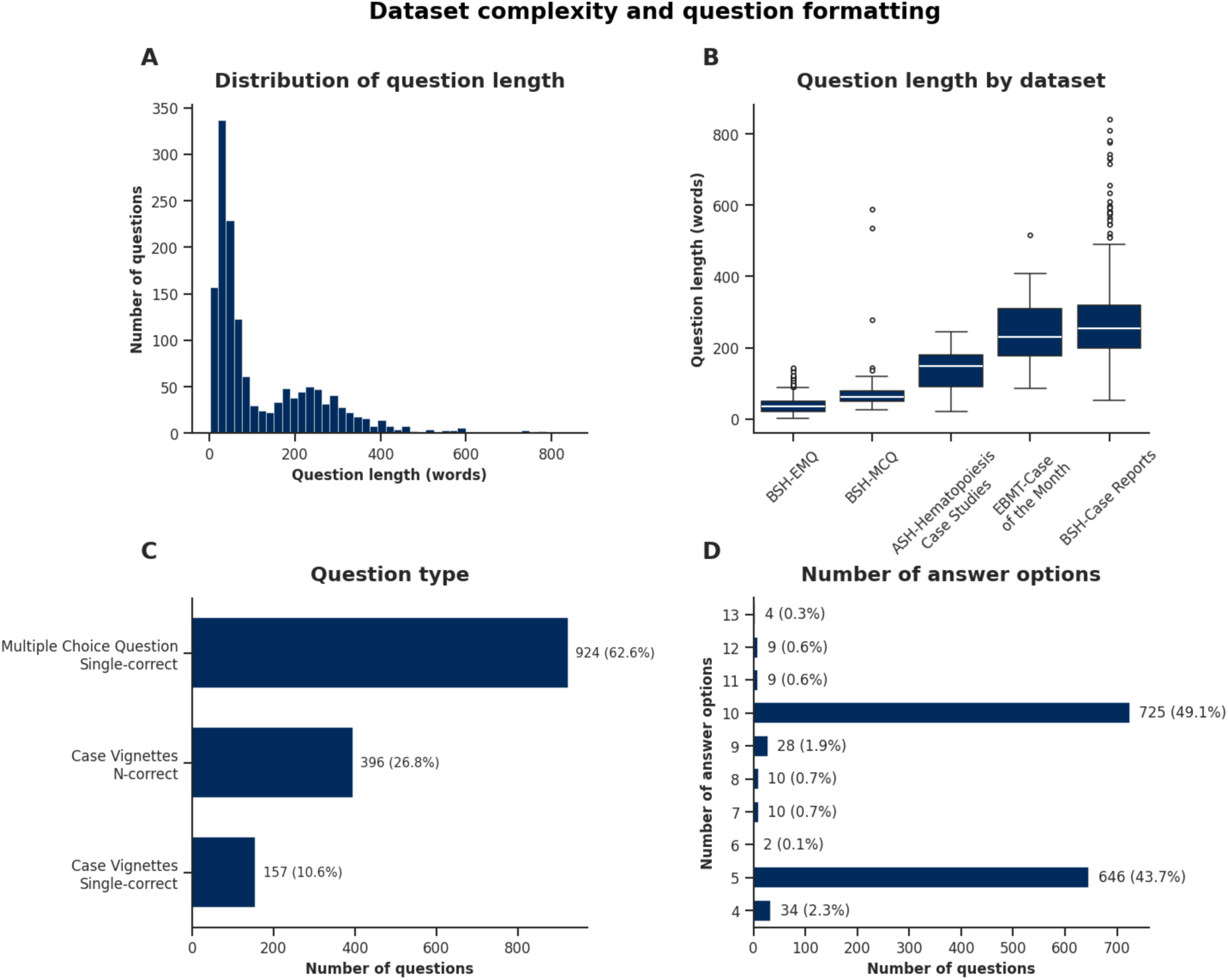
Dataset complexity and question formatting. MCQ length (**A**) followed a right-skewed distribution. Most MCQs contained fewer than 100 words, while a smaller group exceeded 400 words. The median length varied significantly across source datasets (**B**). BSH-EMQ and BSH-MCQ items were typically shorter, whereas EBMT-Case of the Month and BSH-Case Reports featured much longer clinical vignettes, highlighting their increased contextual complexity. The majority of MCQs had a single correct answer option (**C**). In terms of overall answer options, most MCQs had either ten (n=725, 49.1%) or five answer options (n=646, 43.7%; **D**). Labels next to each bar denote the number of questions per category and the percentage of the total dataset in parentheses.

**Fig. S2.**
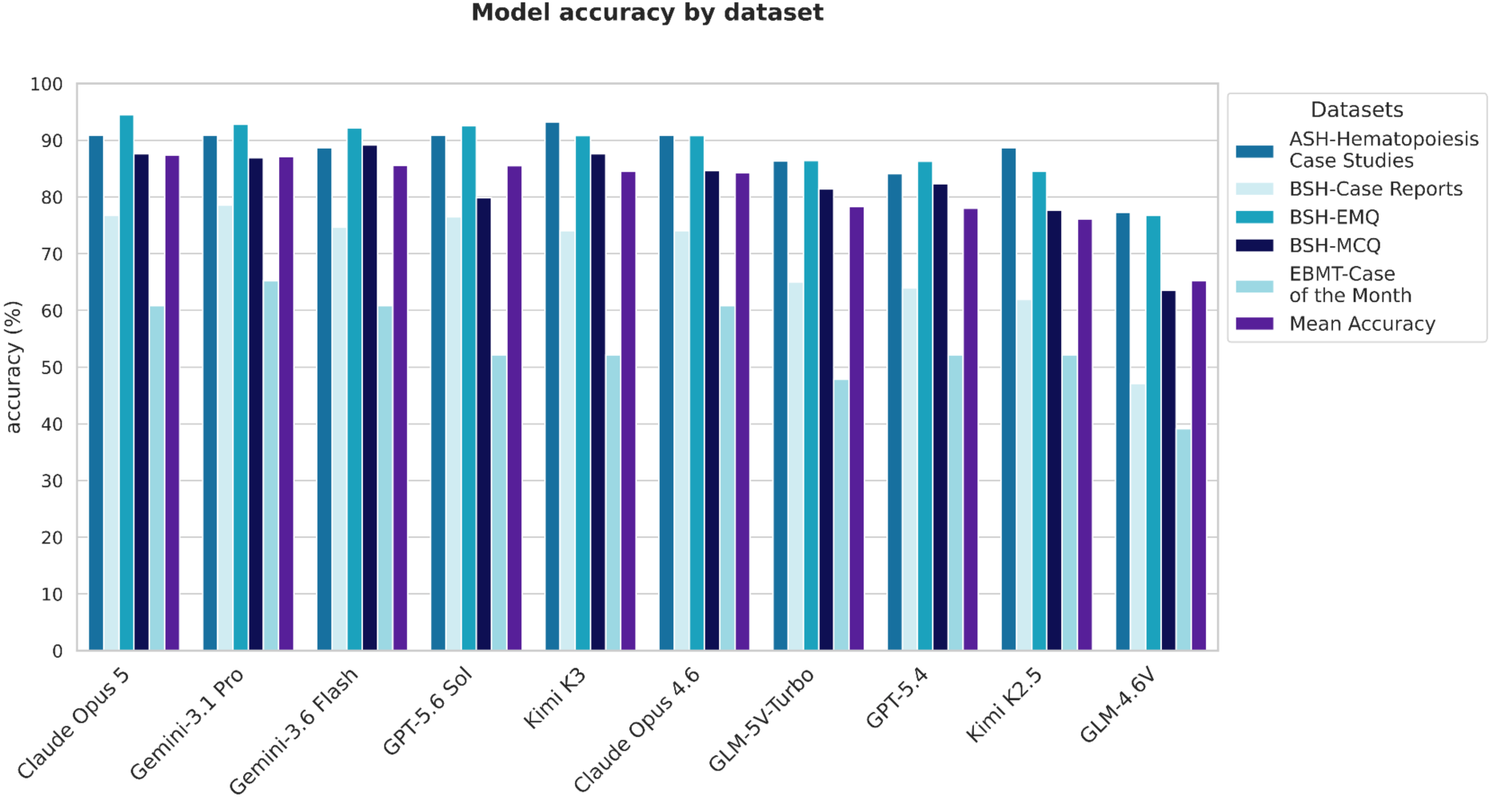
Model accuracy by dataset. Accuracy scores varied between datasets. Models performed better on datasets with only one possible correct answer option (ASH-Hematopoiesis Case Studies, BSH-EMQ and BSH-MCQ) with an average accuracy of 86.3% compared to datasets (EBMT-Case of the Month and BSH-Case Reports) with multiple possible correct answer options, averaging at 61.8%.

**Fig. S3.**
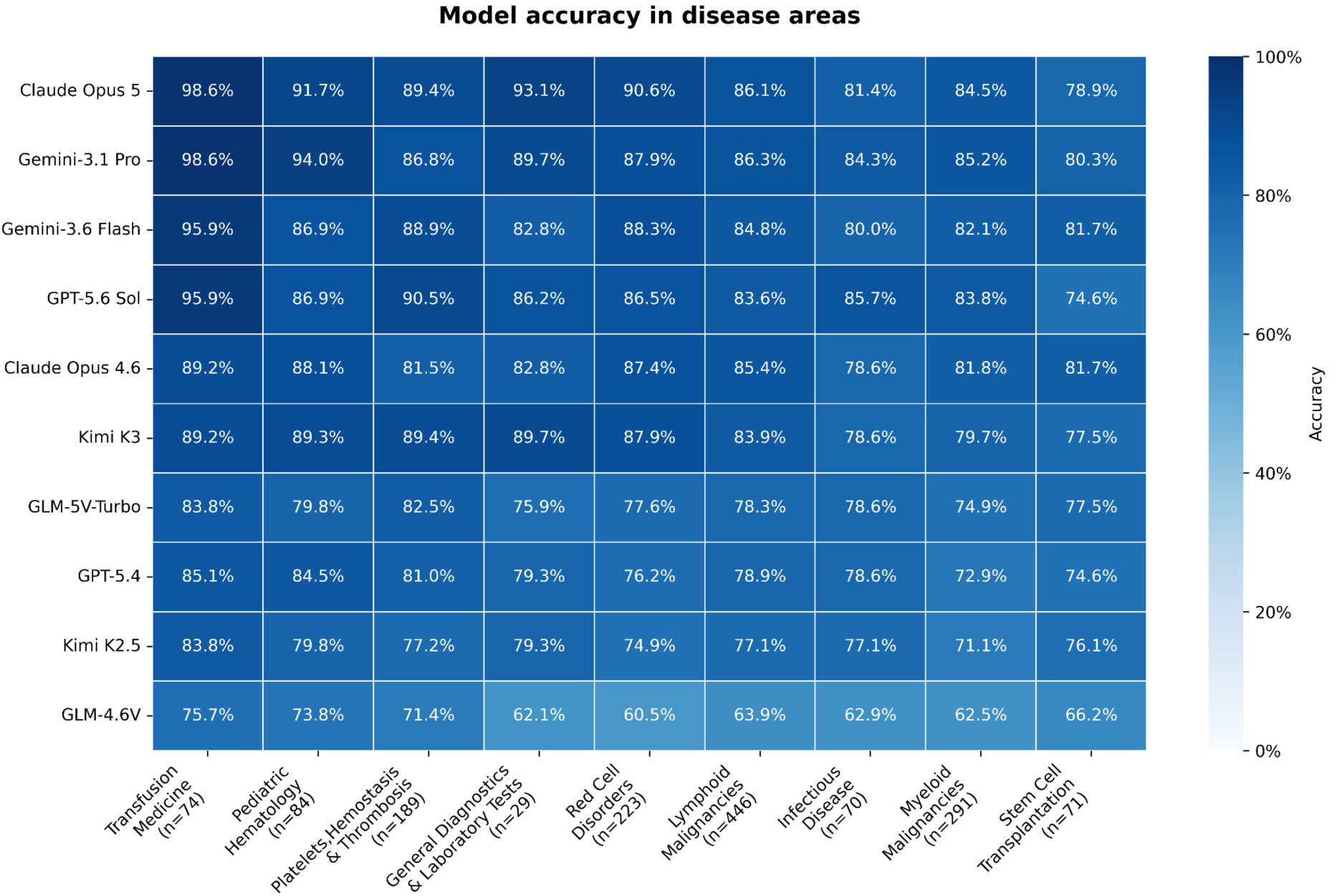
Accuracy results by disease area. The y-axis shows the LLM models, ordered from highest to lowest mean performance (top to bottom). The x-axis displays the disease areas, ordered from best to worst average performance across all models (left to right). For each disease area, the number of questions is given in parentheses.

**Fig. S4.**
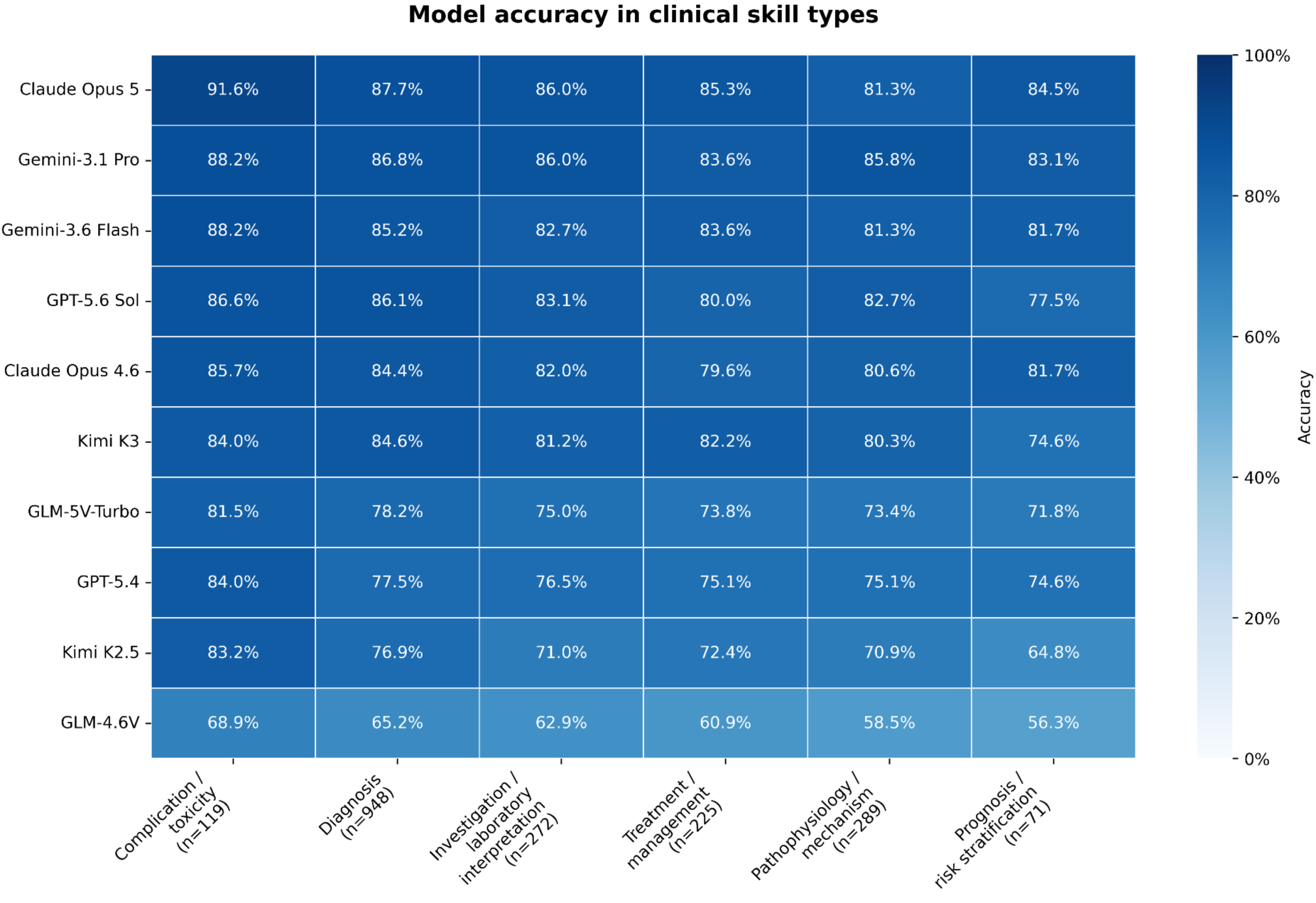
Accuracy results by clinical skill types. The y-axis shows the LLM models, ordered from highest to lowest mean performance (top to bottom). The x-axis displays the clinical skill sets required to solve the cases, ordered from best to worst average performance across all models (left to right). For each clinical skill type, the number of questions is given in parentheses.

**Fig. S5.**
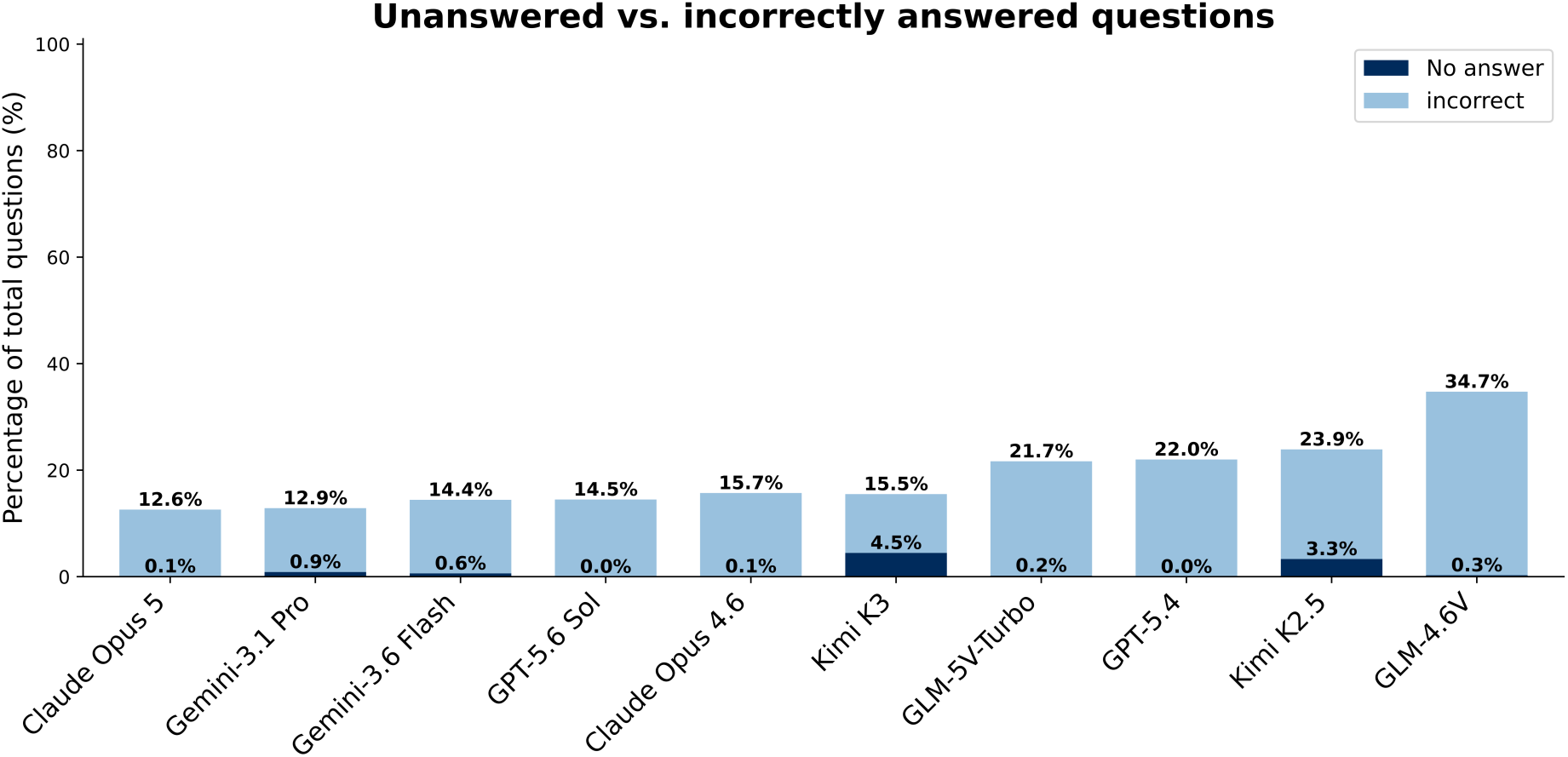
Unanswered vs. incorrectly answered questions. Percentage of total dataset cases which resulted in a model error by either unanswered or incorrectly answered multiple choice questions. This error analysis reveals that generation stalls and token-looping bottlenecks are not a substantial failure indicator in the majority of tested LLMs. Specifically, two of the models produced a correctly formatted response every time, six had a refusal rate under 1% of the MCQs and two models recorded below 4.5% of the total questions as unanswered.

**Fig. S6.**
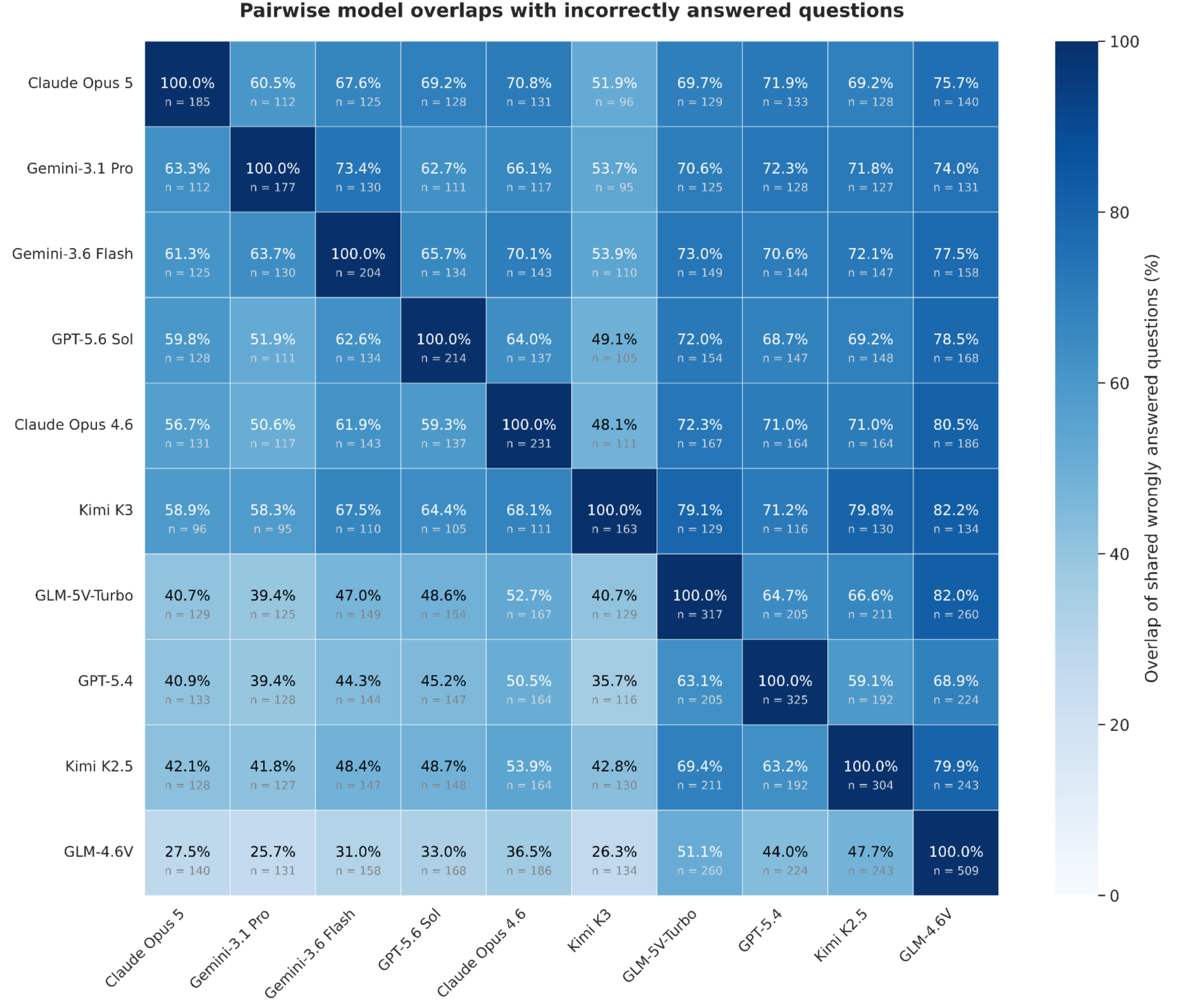
Pairwise model overlaps with incorrectly answered questions. The matrix quantifies the extent to which models fail on the same evaluation questions. Each cell reports the percentage of errors made by the row model that are also incorrect for the column model. The annotations additionally report *n* as the number of shared incorrectly answered questions contributing to each pairwise comparison. The diagonal shows the number of mistakes for each model. Models are ordered by mean accuracy, with higher-performing models positioned toward the top-left of the matrix. The largest absolute overlap values are observed among lower-performing models, which is expected because models with higher error rates have larger error sets and therefore more opportunities for shared mistakes.

